# Data-Driven Hybrid Model of SARIMA-CNNAR For Tuberculosis Incidence Time Series Analysis in Nepal

**DOI:** 10.64898/2026.02.22.26346853

**Authors:** Dip Bahadur Singh, Yashoda Dangi, Pankaj Raj Dawadi

## Abstract

**Background:** Tuberculosis (TB) remains a major public health challenge in Nepal, with incidence rates substantially higher than global estimates. Accurate forecasting of TB incidence is essential for early warning systems, resource allocation, and targeted interventions. This study aimed to develop and validate a hybrid Seasonal Autoregressive Integrated Moving Average (SARIMA) and Convolutional Neural Network Auto-Regressive (CNNAR) model for TB incidence forecasting in Nepal.

**Methods:** Monthly TB incidence data (January 2015 to December 2024) were obtained from the National Tuberculosis Control Center (NTCC), Nepal. A hybrid SARIMA-CNNAR model was developed, where SARIMA modeled linear seasonal trends and CNNAR captured nonlinear patterns in the residuals. Hyperparameters were optimized using grid search with 5-fold cross-validation. Model performance was evaluated using Mean Absolute Error (MAE), Root Mean Square Error (RMSE), Mean Absolute Percentage Error (MAPE), and R^2^ on the 2024 test set. Structural break analysis and sensitivity analysis assessed model robustness. The hybrid model was compared against standalone SARIMA, CNNAR, and three state-of-the-art benchmarks: Long Short-Term Memory (LSTM), Facebook Prophet, and XGBoost.

**Results:** TB incidence in Nepal increased from a monthly average of 2,048 cases in 2015 to 3,447 in 2024 (68.4% increase). The hybrid SARIMA-CNNAR model demonstrated strong performance with test set metrics of MAE=248.35, RMSE=294.31, MAPE=7.2%, and R^2^=0.79. Comparative performance: CNNAR (MAE=251.08, RMSE=336.55, MAPE=7.7%, R^2^=0.73); LSTM (MAE=267.91, RMSE=324.55, MAPE=7.5%, R^2^=0.75); XGBoost (MAE=314.74, RMSE=373.99, MAPE=8.5%, R^2^=0.66); Prophet (MAE=371.15, RMSE=478.40, MAPE=10.4%, R^2^=0.45); SARIMA (MAE=401.11, RMSE=503.93, MAPE=10.99%, R^2^=0.39). All models captured seasonal peaks in March–May and July–August, with forecasts for 2025 indicating continued seasonal patterns. Sensitivity analysis confirmed robustness with <5% metric variation across parameter configurations.

**Conclusions:** This first validated hybrid model for TB prediction in Nepal demonstrates high forecasting accuracy by integrating linear seasonal modeling with nonlinear pattern detection. The approach offers a robust tool for evidence-based public health planning in resource-limited settings and it is suitable for integration into national surveillance systems.

**Author Summary:** Tuberculosis remains a major public health challenge in Nepal, with cases increasing substantially over the past-decade. In this study, we developed a computer model that combines two different forecasting approaches: one that captures regular seasonal patterns and another that learns complex trends from data to predict monthly TB cases. Using ten years of national surveillance data, our hybrid model achieved high accuracy in forecasting TB incidence, outperforming standard approaches including SARIMA, PROPHET, CNNAR, LSTM neural networks, and XGBoost. The model successfully predicted seasonal peaks in March-May and July-August, with forecasts for 2025 suggesting continued high case numbers. These predictions can help Nepal’s health authorities prepare by pre-positioning diagnostic supplies, scheduling additional staffs during peak months, and targeting awareness campaigns. The modeling approach is designed to be adaptable for other diseases and countries with similar health data.

## 1. Introduction

Tuberculosis remains a major global public health challenge, ranking among the top ten causes of death worldwide(1,2). Caused by *Mycobacterium tuberculosis*, the disease manifests in pulmonary and extrapulmonary forms with significant morbidity and mortality burdens(3). According to World Health Organization (WHO) estimates, Nepal faces a disproportionately high TB burden: while approximately 42,000 cases were expected annually, reported new cases reached 69,000, with prevalence and incidence rates 1.8 and 1.6 times higher than estimates, respectively, and mortality 3.1 times higher(4).

Comorbidities such as diabetes substantially increase TB risk and treatment mortality; globally, 371,000 TB cases in 2021 were associated with diabetes, a trend reflected in Nepal’s epidemiological profile(5). Accurate forecasting of TB incidence is essential for effective public health planning, enabling early detection, optimized resource allocation, and targeted interventions(6–8). Time series forecasting models have emerged as valuable tools for understanding disease dynamics and supporting evidence-based decision making(9–11).

Previous studies have employed various modeling approaches for TB prediction across different geographical contexts. Wang et al. developed a SARIMA-NNNAR hybrid model for Qinghai province, China, demonstrating improved accuracy over standalone models(12). Zhao et al. reported over 40% error reduction using an empirical mode decomposition (EMD) ARMA-LSTM hybrid approach(9). Li et al. found that combining ARIMA with Generalized Regression Neural Networks (GRNN) improved short-term TB forecasts in Chinese populations(13,14). Chen et al. applied ARIMA models during the COVID-19 pandemic to forecast TB incidence in Anhui province(15), while Kuan demonstrated the effectiveness of SARIMA and hybrid models in Taiwan(16). Liao et al. assessed trends and predictors of tuberculosis in Taiwan, incorporating demographic and environmental factors(17). Kaur et al. provided a comprehensive review of autoregressive models in environmental forecasting(18), and Li et al. compared time series models with meteorological factors for predicting pulmonary tuberculosis in eastern China(19). Singh et al. conducted epidemiological time series analysis with prediction during the COVID-19 pandemic in Rajasthan, India(20), and Wang et al. developed a SARIMA NARNNX hybrid model for mainland China(21). International applications include TB forecasting in Iran(22), Algeria(6), and assessments of COVID-19 impacts on TB/HIV services in Ghana(23). Zhao et al. compared ARIMA, GM(1,1), and LSTM models for TB prediction in China **[22]**, while Wang et al. proposed a hybrid ARIMA NAR neural network methodology(22). Ragonnet et al. focused on optimizing latency dynamics in TB transmission models(24), and Moosazadeh et al. examined seasonality and temporal variations of tuberculosis in northern Iran(24). Zheng et al. developed forecast models for tuberculosis morbidity in Xinjiang, China(25), and Nepal’s national health sector strategy provides the policy framework for TB control efforts(26).

Despite this growing body of literature, critical gaps remain. First, existing models developed in high-income or middle-income settings may not generalize to low-resource contexts with different epidemiological patterns, health system structures, and data availability. Second, no studies have validated hybrid models specifically for Nepal, which has distinct geographical diversity, population dynamics, and TB burden characteristics. Third, previous approaches often relied on simple feed-forward neural networks without convolutional architectures optimized for time series pattern recognition. Feed-forward networks process inputs in a single direction without feedback loops or memory, limiting their ability to capture sequential dependencies and local patterns in temporal data a limitation that convolutional architectures with autoregressive components can address through hierarchical feature extraction.

The rationale for selecting CNNAR over other deep learning approaches stems from several considerations. Compared to recurrent architectures (LSTM, GRU), CNNAR offers efficient parallelization during training, ability to capture local temporal patterns through convolutional filters, fewer parameters reducing overfitting risk with limited epidemiological time series, and inherent handling of autoregressive structure. Compared to standard feed-forward networks, CNNAR preserves temporal ordering through convolution operations and incorporates autoregressive components specifically designed for time series forecasting. This study aimed to:

1. Develop and validate a data-driven hybrid SARIMA-CNNAR model tailored for time series analysis of tuberculosis incidence in Nepal.
2. Characterize temporal trends, seasonality, and structural breaks in Nepal’s TB incidence data (2015 to 2024).
3. Evaluate forecast accuracy against standalone SARIMA, CNNAR, and three state-of-the-art benchmarks: Long Short-Term Memory (LSTM), Facebook Prophet, and XGBoost.
4. Assess model robustness through comprehensive sensitivity analysis.

## 2. Materials and Methods

### 2.1 Study Design and Data Source

We conducted a retrospective time-series forecasting study using monthly tuberculosis incidence data from Nepal spanning January 2015 to December 2024. Nepal, a South Asian country with high TB burden (estimated incidence 230 per 100,000 population), was selected as the study setting. Aggregated monthly TB incidence data were obtained from the National Tuberculosis Control Center (NTCC), Nepal, extracted via the District Health Information System 2 (DHIS2) platform. The dataset comprised district-wise case counts aggregated to the national level, covering all seven provinces and three ecological regions over 120 months (January 2015 to December 2024). Data underwent institutional validation at NTCC prior to release. Total observations: 120 monthly time points (108 months for training, 12 months for testing).

### 2.2 Ethical Considerations

Ethical approval was obtained from the Nepal Health Research Council (NHRC) (Protocol No. 547-2024, dated December 24, 2024). The study adhered to NHRC guidelines, WHO standards, and the Declaration of Helsinki. All data were fully anonymized with no personally identifiable information; district-level aggregates were processed without individual identifiers. Data security protocols ensured confidential handling and prevention of misuse.

### 2.3 Data Preprocessing

Missing values (2.4% of monthly entries) were identified using pivot-based detection and imputed using Kalman filtering, which maintains time-series integrity by modeling the underlying state-space structure. Kalman filtering was selected over mean imputation or interpolation due to its ability to incorporate temporal dependencies and measurement uncertainty.

Outliers were identified using the Interquartile Range (IQR) method, defined as values below Q1-1.5×IQR or above Q3 + 1.5×IQR. Detected outliers (n=6) were examined for data entry errors versus genuine epidemiological signals. True outliers were capped at the adjacent non-outlier bounds; erroneous entries were corrected against source records.

Nepali calendar dates (Bikram Sambat) were converted to Gregorian calendar dates for international standardization and alignment with external datasets (Gregorian Year = Nepali Year-57). For neural network components, data were normalized to [0,1] range using min-max scaling:

Xnorm 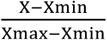. Normalization parameters were derived solely from the training set and applied to test data to prevent data leakage.

### 2.4 Exploratory data analysis

Comprehensive EDA was conducted to characterize data structure and inform model development: Time trend analysis: Temporal plots with LOESS smoothing to visualize overall trend, seasonality, and anomalies.

Summary statistics: Monthly and annual means, medians, standard deviations, IQRs, and ranges stratified by year.

Seasonal decomposition: Additive decomposition using STL (Seasonal-Trend decomposition using LOESS) to separate observed series into trend, seasonal, and residual components.

Correlation analysis: Pearson correlation matrices examining relationships between years; autocorrelation function (ACF) and partial autocorrelation function (PACF) plots to identify temporal dependencies.

Structural break detection: Bai-Perron multiple breakpoint test to identify significant shifts in the time-series mean or trend.

Distribution analysis: Histograms, Q-Q plots, and boxplots to assess normality, skewness, and outlier presence before and after preprocessing.

### 2.5 Stationarity assessment

The Augmented Dickey–Fuller (ADF) test is used to assess stationarity by testing for the presence of a unit root in a time series. The test is based on the regression model

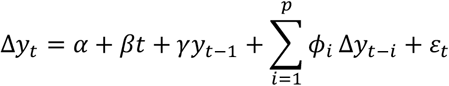

where *y*_*t*_is the observed series, Δ*y*_*t*_ = *y*_*t*_ − *y*_*t*−1_is the first difference, *α*is a constant, *βt*is an optional trend term, *γ*measures the effect of the lagged level, *ϕ*_*i*_are coefficients of lagged differences, and *ε*_*t*_is the error term. The null hypothesis (*H*_0_) states that the series has a unit root (non-stationary), while the alternative hypothesis (*H*_*a*_) states that it is stationary. A p-value greater than 0.05 indicates non-stationarity, whereas a p-value less than or equal to 0.05 indicates stationarity. The test therefore determines whether the series exhibits mean-reverting behavior or unit root characteristics.

### 2.6 SARIMA Model Specification

The Seasonal Autoregressive Integrated Moving Average (SARIMA) model extends ARIMA to accommodate seasonality in time-series data. The general multiplicative form of the SARIMA model is:

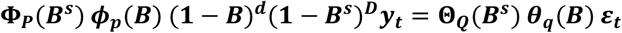

#### Parameters

p (non-seasonal AR order), d (non-seasonal differencing), q (non-seasonal MA order), P (seasonal AR order), D (seasonal differencing), Q (seasonal MA order), s (seasonal period, s=12 for monthly data), B (backshift operator), *ε*_*t*_ (white noise error).

#### Model selection

Candidate models were evaluated using Akaike Information Criterion (AIC) and Bayesian Information Criterion (BIC). Grid search over p,d,q ∈ {1,1,1} and P,D,Q ∈ {1,1,1} with s=12 was conducted. Stationarity and invertibility were verified. The optimal model SARIMA (1,1,1)(1,1,1)[12] minimized AIC (2,847.3) and BIC (2,872.6).

### 2.7 CNNAR Model Architecture

The Convolutional Neural Network Auto-Regressive (CNNAR) model is designed to capture nonlinear patterns in time series data through convolutional feature extraction. Mathematically, the model is expressed as 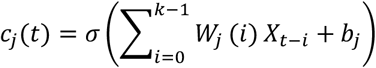

The equation represents a part of the model’s computation, where *c*_*j*_(*t*)is the output at time t for the j-th feature. It is calculated by an activation function σ (commonly a ReLU or sigmoid function) to the weighted sum of the input features, where W_j_(i) are the weights corresponding to the lagged observations X(t−i) for i=1 to K-1, and b_j_ is the bias term. The equation models the interaction between the current time step and its past values, capturing the temporal dependencies in the data.

Fig1. Prediction intervals for the CNNAR model were generated using a bootstrap resampling approach(27). Specifically, after training the CNNAR on the full training set, residuals from the training predictions were computed. One thousand bootstrap samples were then generated by randomly sampling these residuals with replacement and adding them to the point forecasts, producing an empirical distribution of possible future values. The 2.5th and 97.5th percentiles of this bootstrap distribution were used as the 95% prediction intervals. This non-parametric approach was chosen because it makes no assumptions about the distribution of forecast errors and captures the heteroskedasticity observed in the residuals (ARCH test p=0.0039, Table 8).

**Fig 1.**
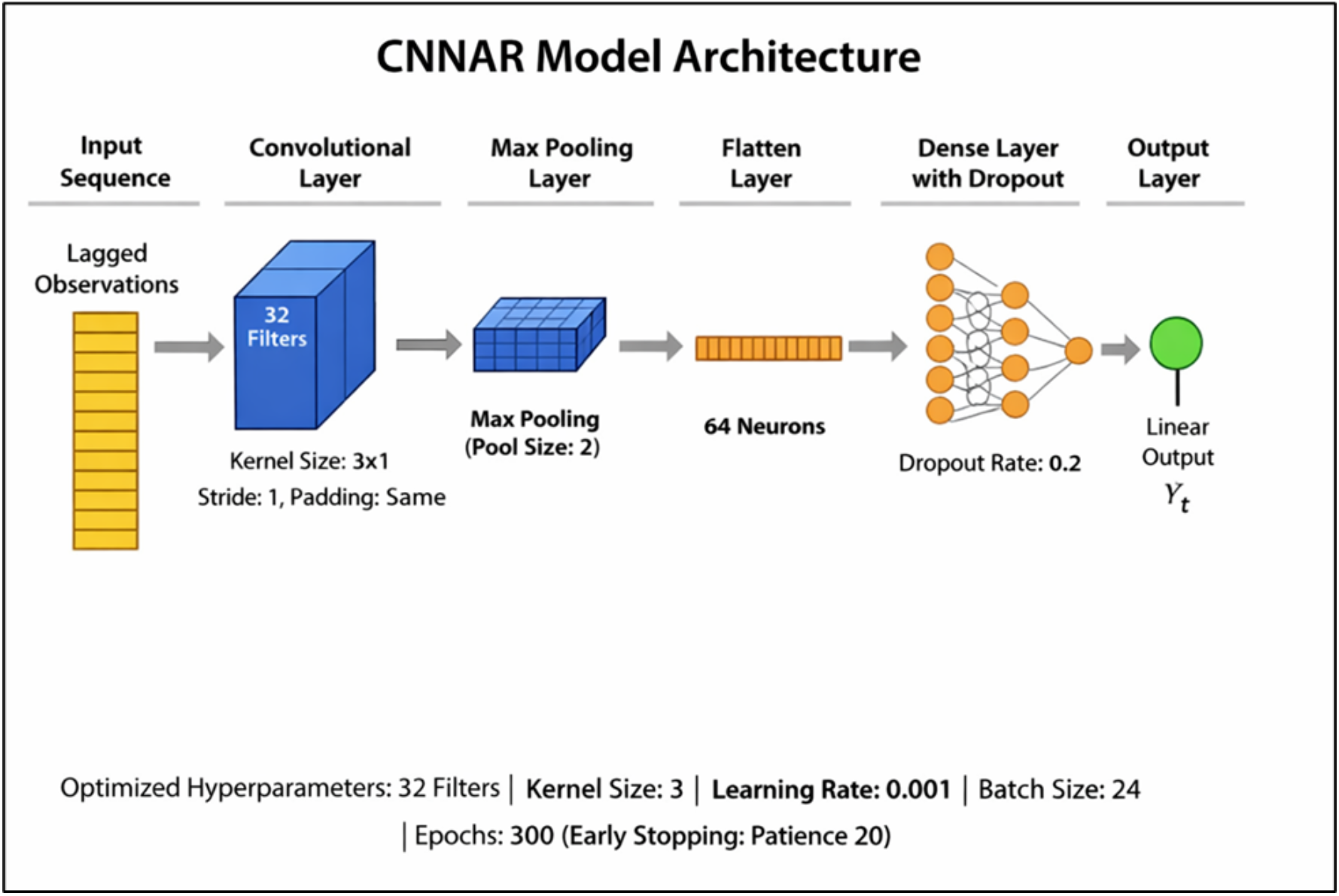
CNNAR Model Architecture. Two convolutional layers (64 and 32 filters) with ReLU activation, dropout (0.2), and dense output layer for one-step-ahead forecasting.

### 2.8 Hybrid SARIMA-CNNAR framework

The hybrid model combines linear predictions from SARIMA with nonlinear residual modeling by CNNAR, the hybrid prediction is expressed as: 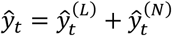.

Where 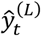 represents the linear and seasonal component captured by SARIMA and 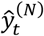 represents the nonlinear component modeled by CNNAR.

Equivalently, the hybrid formulation can be written as: *ŷ*_*t*_ = SARIMA(*y*_*t*_) + CNNAR(*y*_*t*_ − SARIMA(*y*_*t*_)). The meaning that CNNAR is applied to the residuals obtained after fitting the SARIMA model. The complete mathematical representation is:*ŷ*_*t*_ = Φ_*P*_(*B*^*s*^)*ϕ*_*p*+_(*B*)(1 − *B*)^*d*^ (1 − *B*^*s*^)^*D*^ *y*_*t*_ + *f*(*σ*(*W*_*c*_ ∗ *E*_*t*_ + *b*_*c*_)),

Where *ŷ*_*t*_ denotes the final predicted value, Φ_*P*_(*B*^*s*^)*ϕ*_*p*+_(*B*)(1 − *B*)^*d*^ (1 − *B*^*s*^)^*D*^ *y*_*t*_ represents the seasonal and non-seasonal SARIMA operators, and *f*(*σ*(*W*_*c*_ ∗ *E*_*t*_ + *b*_*c*_)) denotes the CNNAR nonlinear mapping. In this formulation, *E*_*t*_ = [*e*_*t*−1_, *e*_*t*−2_, . . ., *e*_*t*−*p*_] are the lagged residuals from SARIMA, *W*_*c*_is the convolutional filter weight matrix, ∗denotes the convolution operation, *b*_*c*_is the bias term, *σ*(⋅)is the ReLU activation function, and *f*(⋅)represents the dense layer transforming extracted features into the nonlinear forecast component.

Fig2. generates prediction intervals for the hybrid model, we combined the uncertainty from both components. First, 1,000 bootstrap samples of the CNNAR residual forecasts were generated as described in Section 2.7. Second, the SARIMA model’s forecast variance was obtained from its parametric covariance matrix. The total forecast variance for each time point was calculated as the sum of the SARIMA variance and the bootstrap variance of the CNNAR component, assuming independence between the two sources of uncertainty(28). The 95% prediction intervals were then constructed as the point forecast ± 1.96 × √(total variance). This approach acknowledges that uncertainty propagates from both the linear and nonlinear components of the hybrid model.

**Fig 2.**
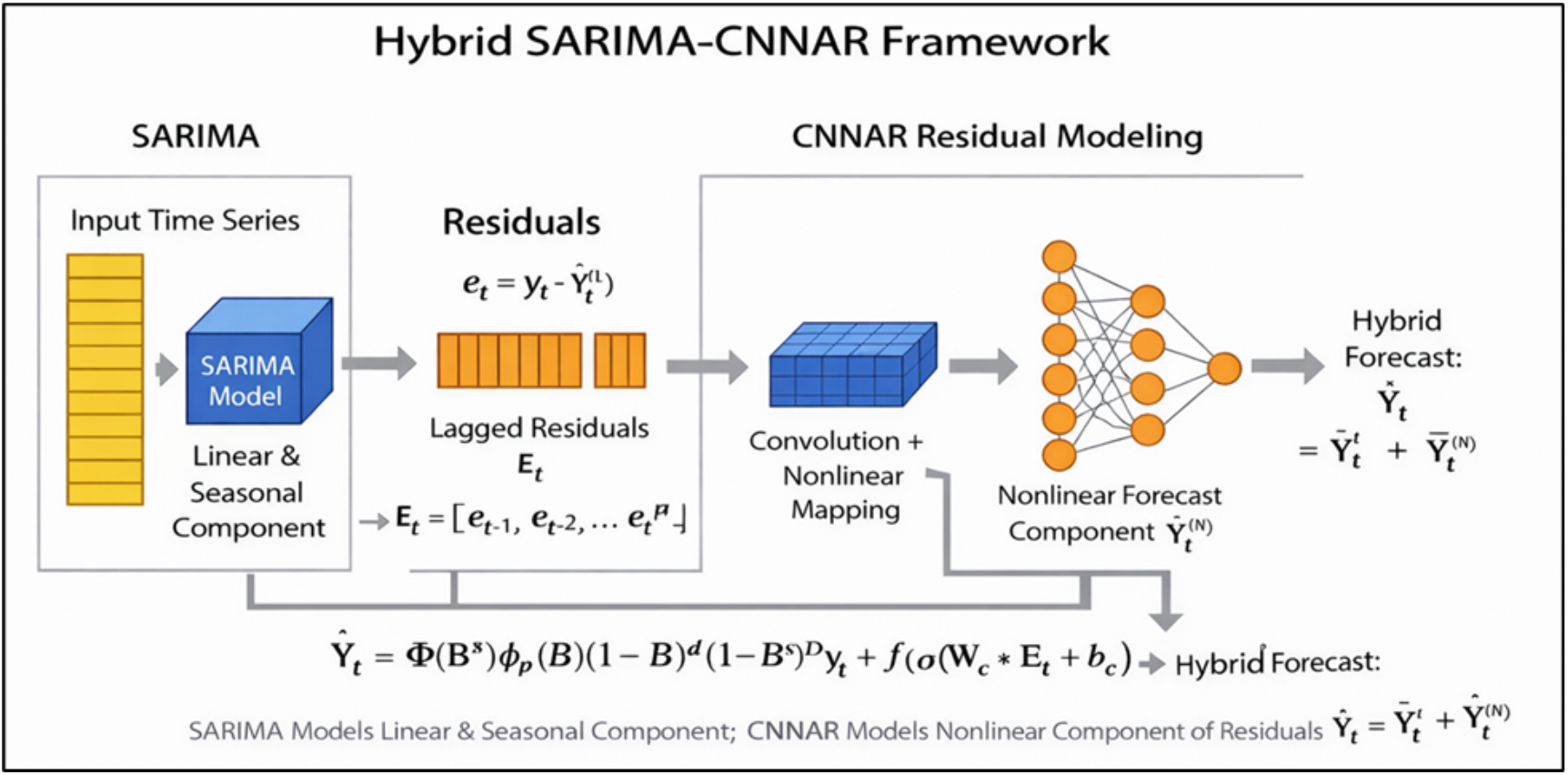
Hybrid SARIMA-CNNAR Framework. Two-stage process: SARIMA captures linear trend and seasonality; CNNAR models nonlinear patterns in residuals. Final forecast combines both components.

#### Residual analysis

was conducted on SARIMA residuals to justify the hybrid approach. The Ljung–Box test was applied to assess remaining autocorrelation, and the BDS test was used to detect nonlinearity. The presence of statistically significant nonlinear patterns (p < 0.01) confirmed that SARIMA alone was insufficient, thereby supporting the application of the CNNAR model to capture additional nonlinear structure in the residual series.

### 2.9 Benchmark Models write the mathematical equation

To contextualize performance regarding state-of-the-art comparison, we evaluated three additional forecasting models alongside SARIMA and CNNAR.

#### 2.9.1 Long Short-Term Memory (LSTM)

Let the input sequence consist of 12 lagged observations:

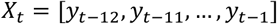

The LSTM cell dynamics for each time step are:

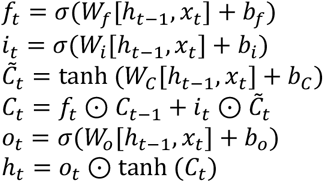

The stacked architecture is represented as:

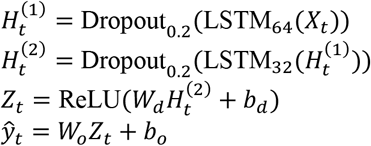

The LSTM captures nonlinear temporal dependencies through gated memory cells. Two stacked LSTM layers (64 and 32 units) extract hierarchical temporal features, followed by a dense transformation for one-step-ahead forecasting. Optimization is performed using Adam with learning rate 0.001.

#### 2.9.2 Facebook Prophet

Prophet uses an additive decomposition:

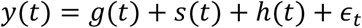

Seasonality via Fourier series:

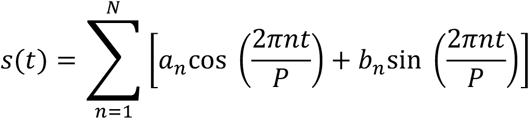

where *P* = 12.

Prophet decomposes time series into trend *g*(*t*), seasonality *s*(*t*), holiday effects *h*(*t*), and error. Fourier terms allow flexible seasonal patterns.

#### 2.9.3 XGBoost

The objective at iteration *t*:

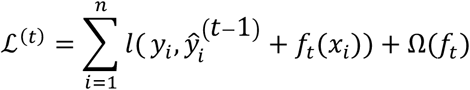

Regularization:

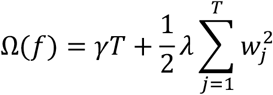

XGBoost minimizes prediction error with regularization to control model complexity. *T*is the number of leaves, and *γ, λ*penalize overfitting.

### 2.10 Evaluation Metrics

Forecasting performance was assessed using four standard evaluation metrics: Mean Absolute Error (MAE), Root Mean Square Error (RMSE), Mean Absolute Percentage Error (MAPE), and the Coefficient of Determination (R^2^).

The Mean Absolute Error (MAE) measures the average magnitude of forecast errors and is defined as

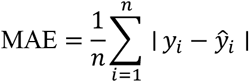

where *y*_*i*_ is the actual value and *ŷ*_*i*_is the predicted value.

The Root Mean Square Error (RMSE) evaluates the square root of the average squared prediction errors and is given by

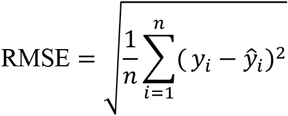

RMSE penalizes larger errors more heavily due to squaring.

The Mean Absolute Percentage Error (MAPE) expresses forecast accuracy as a percentage and is calculated as

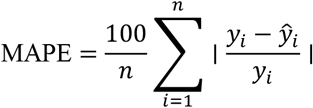

This metric provides scale-independent interpretability.

The Coefficient of Determination (R^2^) measures the proportion of variance explained by the model and is defined as

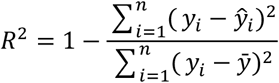

where 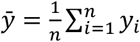 represents the sample mean of the observed values.

Together, these metrics provide a comprehensive assessment of forecast accuracy, error magnitude, percentage deviation, and explanatory power.

### 2.11 Model validation

Data splitting: The 120-month dataset was divided into two parts: the training set, which spans from January 2015 to December 2023, covering 108 months or 90% of the data, and the test set, which includes the remaining 12 months, from January 2024 to December 2024, making up 10% of the dataset.

Cross-validation: Five-fold time-series cross-validation was employed for hyperparameter tuning, preserving temporal order in each fold.

Statistical tests on residuals: Normality (Shapiro-Wilk test), autocorrelation (Ljung-Box test), and heteroskedasticity (ARCH test) were assessed.

Structural break incorporation: Identified breaks were assessed through sensitivity analysis rather than explicit modeling to avoid overfitting.

### 2.12 Sensitivity analysis

The sensitivity analysis involved varying key parameters for both SARIMA and CNNAR models. For SARIMA, parameters (p, d, q, P, D, Q) were adjusted by ±1 from optimal values, and seasonal periods of 11, 12, and 13 were tested. For CNNAR, hyperparameters like the number of filters (16, 32, 64, 128), kernel size (2, 3, 4), learning rate (0.0005, 0.001, 0.005), and dropout rate (0.1, 0.2, 0.3) were varied. Training configurations, including window lengths (96, 102, 108 months) and the inclusion/exclusion of the COVID-19 period (2020-2021), were also tested. Performance variation across configurations was quantified using the coefficient of variation (CV) for each metric.

### 2.13 Software implementation

All analyses were performed in Python 3.12.7 using: The analysis in this study utilized several Python libraries to perform various tasks. Statsmodels (v0.14.0) was used for SARIMA modeling, conducting Augmented Dickey-Fuller (ADF) tests, and time series decomposition. For implementing the CNNAR model, TensorFlow/Keras (v2.13.0) was employed. Scikit-learn (v1.3.0) played a crucial role in data preprocessing, performing cross-validation, and calculating evaluation metrics. Data manipulation tasks were handled with Pandas (v2.1.0), while NumPy (v1.24.0) was used for efficient numerical computations. To visualize the results, Matplotlib/Seaborn (v3.7.0) were used for creating plots and charts. Arch (v6.0) was employed for heteroskedasticity testing, and Ruptures (v1.1.0) was used to detect structural breaks in the time series data.

## 3. Results

### 3.1 Descriptive epidemiology

Table 1 shows the aggregated monthly TB incidence data were obtained from the National Tuberculosis Control Center (NTCC), Nepal, extracted via the District Health Information System 2 (DHIS2) platform. The dataset comprised district-wise case counts aggregated to national level, covering all seven provinces and three ecological regions over 120 months (January 2015 to December 2024). Data underwent institutional validation at NTCC prior to release. Total observations: 120 monthly time points (108 months for training, 12 months for testing).

**Table 1.** Complete monthly TB incidence dataset (2015-2024). De-identified monthly tuberculosis case counts in Nepal from January 2015 to December 2024. Source: National Tuberculosis Control Center (NTCC).

| Month |  | 2015 | 2016 | 2017 | 2018 | 2019 | 2020 | 2021 | 2022 | 2023 | 2024 |
| --- | --- | --- | --- | --- | --- | --- | --- | --- | --- | --- | --- |
| 1 | January | 1410 | 1533 | 1667 | 1812 | 2008 | 2475 | 2293 | 2292 | 2599 | 2991 |
| 2 | February | 1986 | 2117 | 2257 | 2406 | 2396 | 2675 | 2739 | 3208 | 2977 | 3531 |
| 3 | March | 2147 | 2311 | 2488 | 2679 | 2627 | 1881 | 3370 | 3775 | 3620 | 4173 |
| 4 | April | 2302 | 2457 | 2622 | 2798 | 2979 | 1772 | 2450 | 3269 | 3483 | 4132 |
| 5 | May | 2217 | 2407 | 2612 | 2835 | 3043 | 2069 | 1562 | 3835 | 4150 | 4635 |
| 6 | June | 2400 | 2531 | 2668 | 2813 | 2735 | 2472 | 2475 | 3669 | 3453 | 3863 |
| 7 | July | 2287 | 2422 | 2566 | 2718 | 2676 | 2782 | 3503 | 3562 | 3608 | 3839 |
| 8 | August | 2242 | 2323 | 2406 | 2493 | 2692 | 2093 | 3136 | 3197 | 2982 | 3082 |
| 9 | September | 2174 | 2217 | 2260 | 2305 | 2030 | 2500 | 2572 | 2480 | 3037 | 2592 |
| 10 | October | 1878 | 1948 | 2022 | 2098 | 2145 | 1865 | 2593 | 2613 | 2027 | 2619 |
| 11 | November | 1741 | 1851 | 1969 | 2094 | 2400 | 2016 | 2992 | 2789 | 3012 | 3030 |
| 12 | December | 1790 | 1887 | 1990 | 2098 | 2255 | 2457 | 2944 | 2524 | 2784 | 2882 |

Table 2 shows the TB incidence in Nepal increased from a monthly average of 2,048 cases (SD=292) in 2015 to 3,447 cases (SD=674) in 2024, representing a 68.4% increase over the decade. The highest recorded monthly incidence was 4,635 cases (May 2024); the lowest was 1,410 cases (January 2015). Distinct seasonal patterns emerged with consistent peaks in March-May and July-August, and troughs in February and September.

**Table 2.**
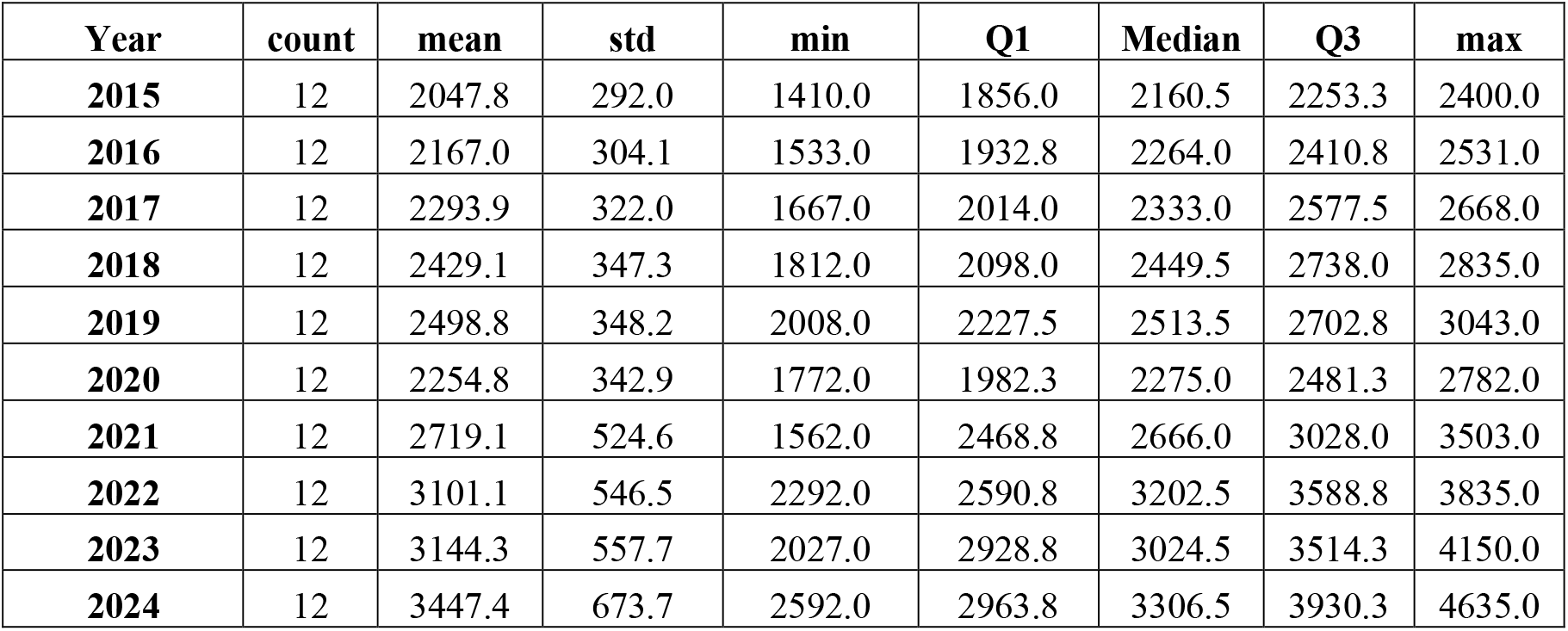
Annual Descriptive Statistics of TB Incidence (2015-2024). Summary statistics by year showing 68.4% increase in mean monthly cases from 2,048 (2015) to 3,447 (2024).

### 3.2 Trend analysis

Fig3. Monthly TB incidence in Nepal (2015-2024). The plot shows clear seasonal fluctuations with periodic peaks and troughs, and a notable overall increase after 2020. The blue line represents monthly cases; the red line shows LOESS smoothing.

**Fig 3.**
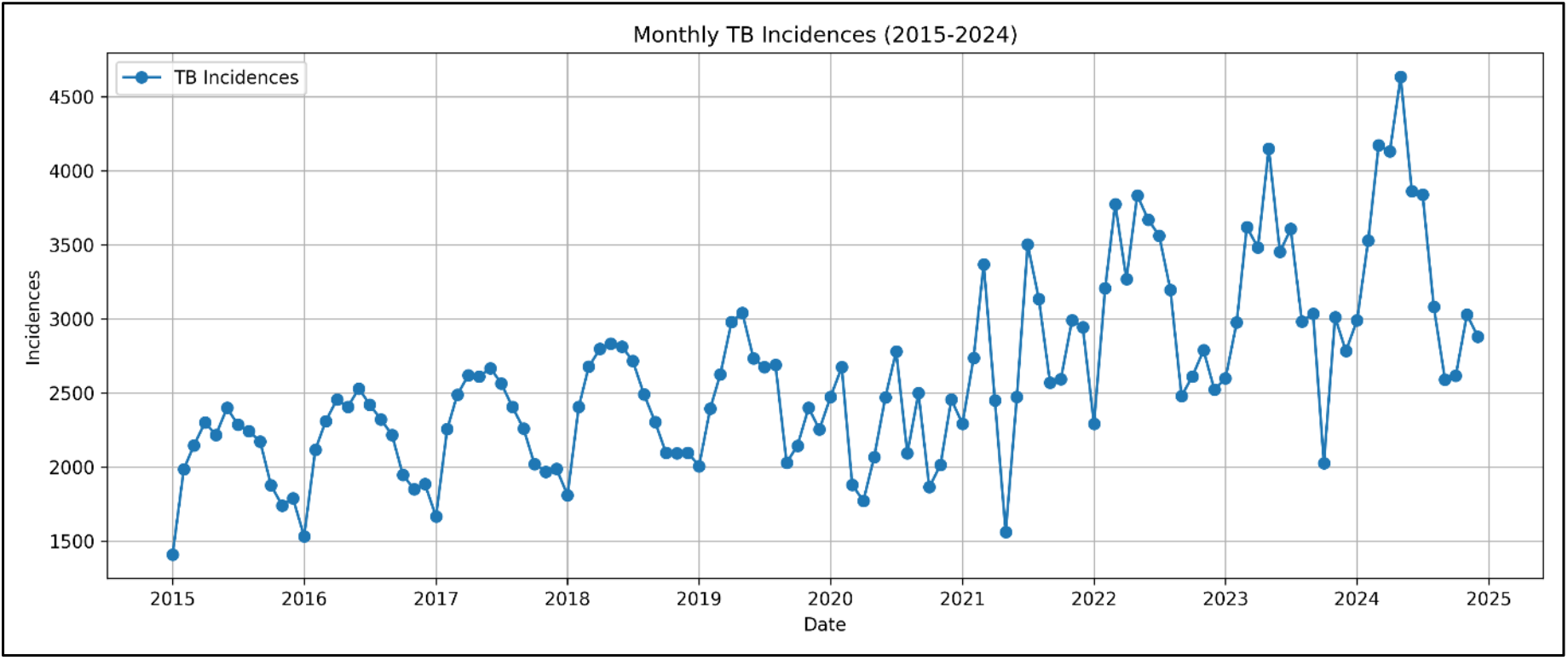
Monthly TB incidence trend with LOESS smoothing. Blue line: monthly cases; red line: LOESS smoothing. Shows seasonal fluctuations and post-2020 increase.

The temporal dynamics reveal: The TB incidence data shows distinct phases. During the pre-pandemic period (2015-2019), there was stable seasonality with a 4.5% annual increase. In 2020, the pandemic caused a sharp decline, with April’s cases falling 42% below the 2019 average, likely due to reduced healthcare-seeking and TB detection. The recovery phase (2021-2022) saw a gradual return to pre-pandemic trends, surpassing previous levels by 2022. In the post-pandemic period (2023-2024), TB incidence remained elevated, consistently exceeding 3,000 cases monthly, with multiple months over 4,000.

### 3.3 Distributional characteristics

Fig4. Histogram of TB incidences with density curve. The distribution peaks around 2,500 cases with slight right-skewness (skewness=0.84). The density curve confirms concentration near the central value with gradual decline at higher levels.

**Fig 4.**
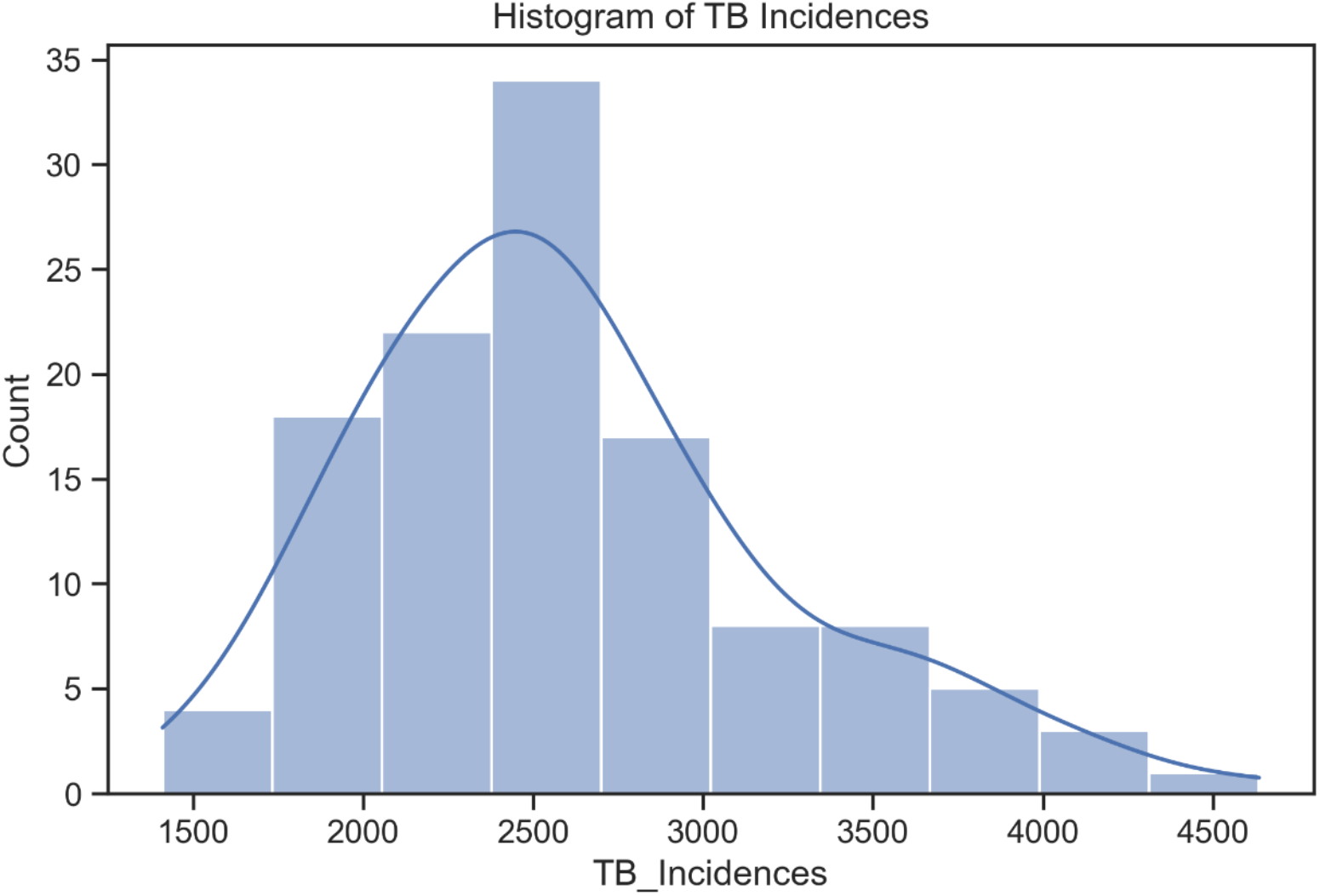
Distributional characteristics. Histogram with density curve. Distribution peaks at ∼2,500 cases with slight right-skewness (skewness=0.84).

Fig5. Q-Q plot for normality assessment. Points generally align with the reference line, suggesting approximate normality. Minor deviations at the upper end indicate some extreme values. Formal normality testing using the Shapiro-Wilk test on the full series yielded W=0.97, p=0.08, failing to reject normality at α=0.05.

**Fig 5.**
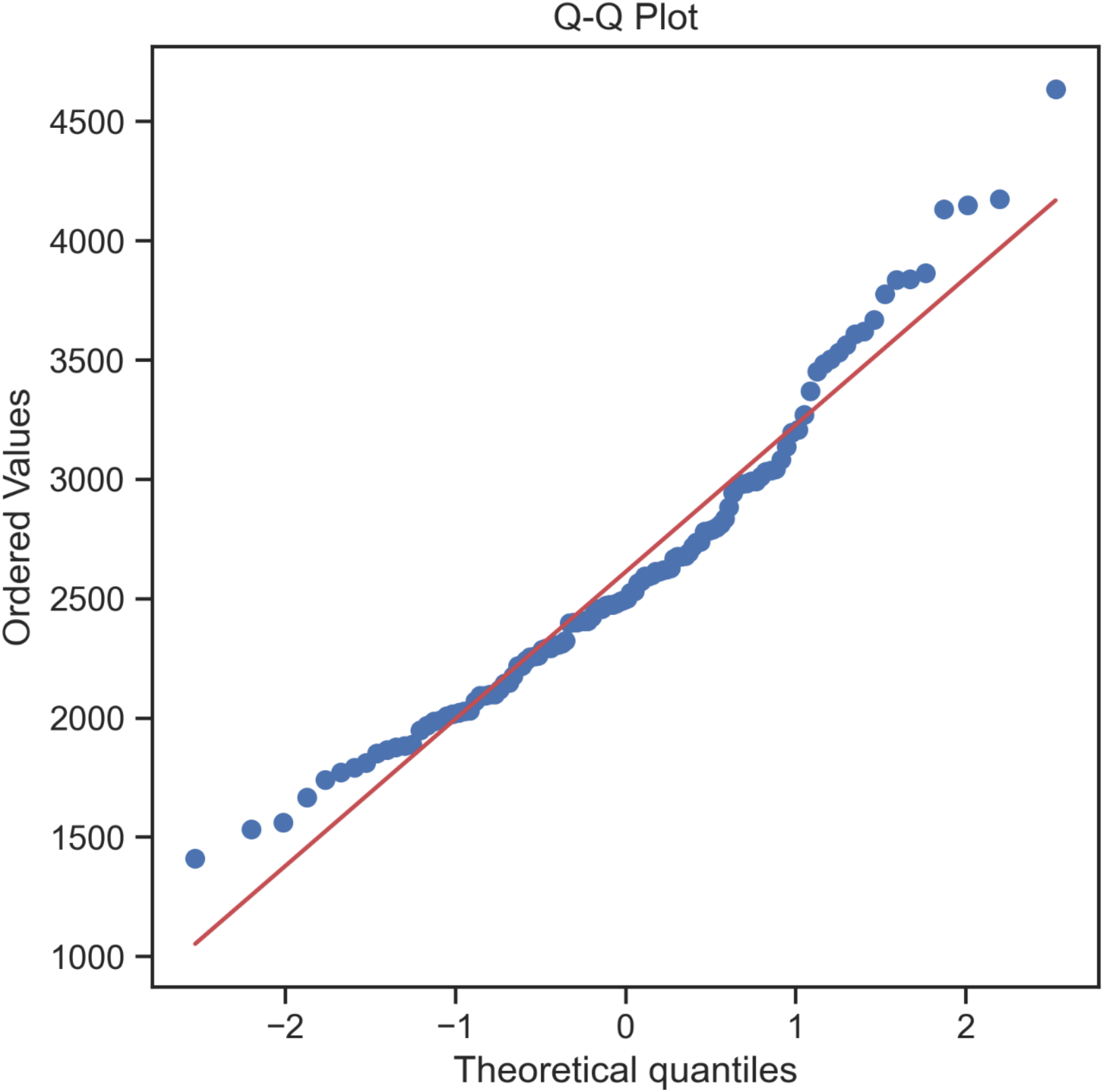
Q–Q Plot for Normality Assessment. Points align with reference line (W=0.97, p=0.08), suggesting approximate normality. Minor deviations at upper end.

### 3.4 Correlation structure

Fig6. Correlation heatmap of annual TB incidences. Most years show strong positive correlations (r>0.7), with 2020 showing notably lower correlations reflecting pandemic disruption. Values range from -0.5 (blue) to 1.0 (red). Key findings from the analysis reveal strong positive correlations (r > 0.7) between consecutive years, especially from 2015 to 2019. The highest correlation was observed between 2018 and 2019 (r = 0.84, p < 0.001). However, in 2020, correlations were notably lower, including negative correlations with 2019 (r = -0.32) and 2023 (r = -0.42), reflecting the disruption caused by the pandemic. After 2020, correlations strengthened again, with the 2022-2023 period showing a strong correlation of r = 0.76.

**Fig 6.**
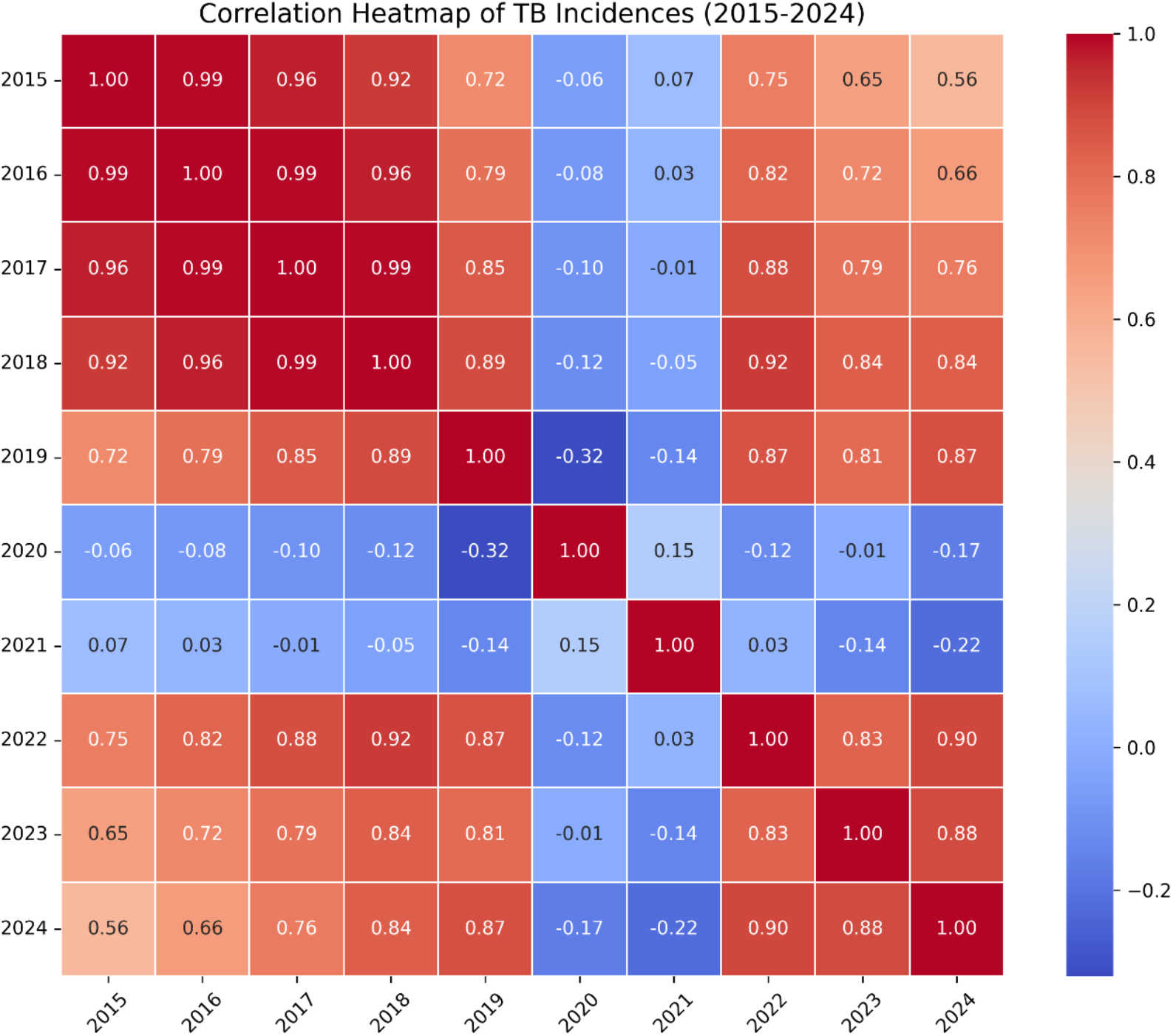
Correlation Heatmap of Annual TB Incidences. Strong positive correlations (r>0.7) between most years; 2020 shows lower correlations reflecting pandemic disruption.

### 3.5 Outlier analysis and treatment

Fig7. Boxplot comparison before and after outlier treatment. The “After” panel shows capped extreme values while preserving the upward trend and distributional shape.

**Fig 7.**
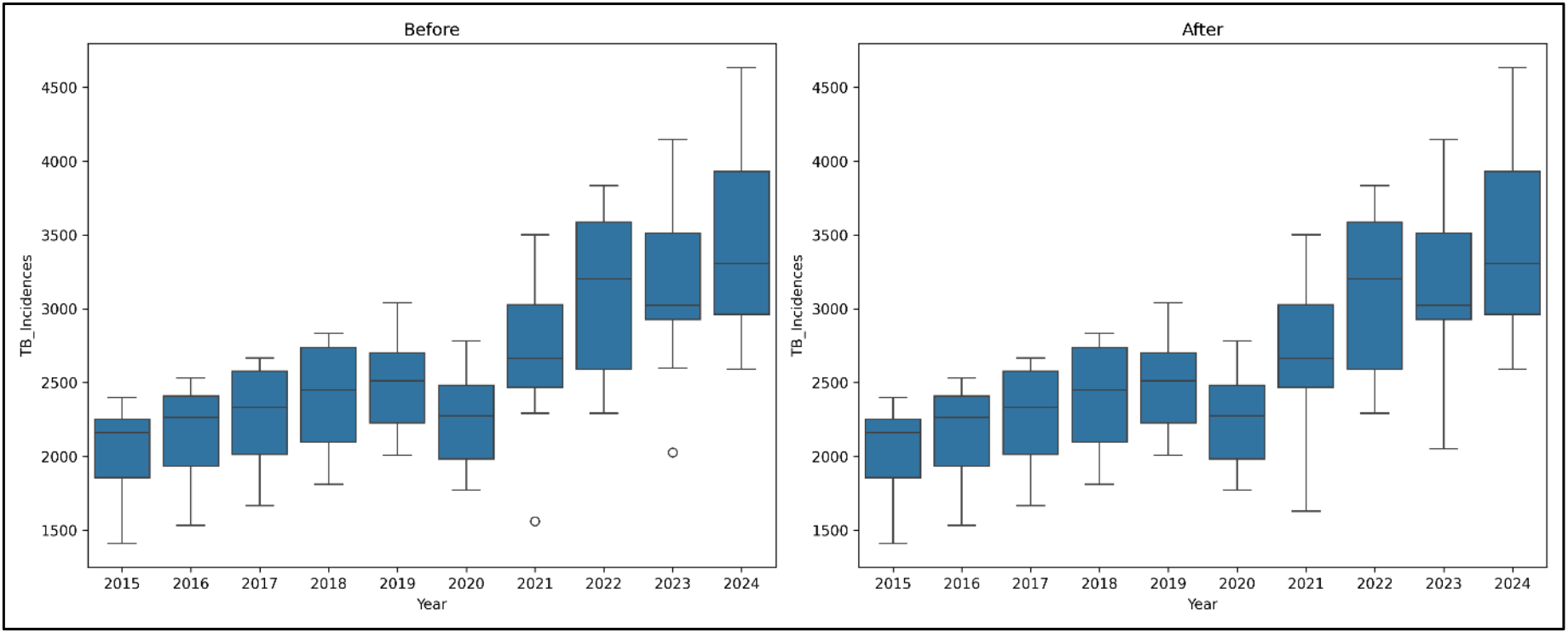
Outlier analysis and treatment. Boxplot comparison before/after treatment. Six outliers identified via IQR method were capped at adjacent bounds.

### 3.6 Autocorrelation structure

Fig8. ACF and PACF plots. (A) ACF shows significant correlations at lags 1, 2, and 12. (B) PACF shows sharp spike at lag 1 and significant spike at lag 12.

**Fig 8.**
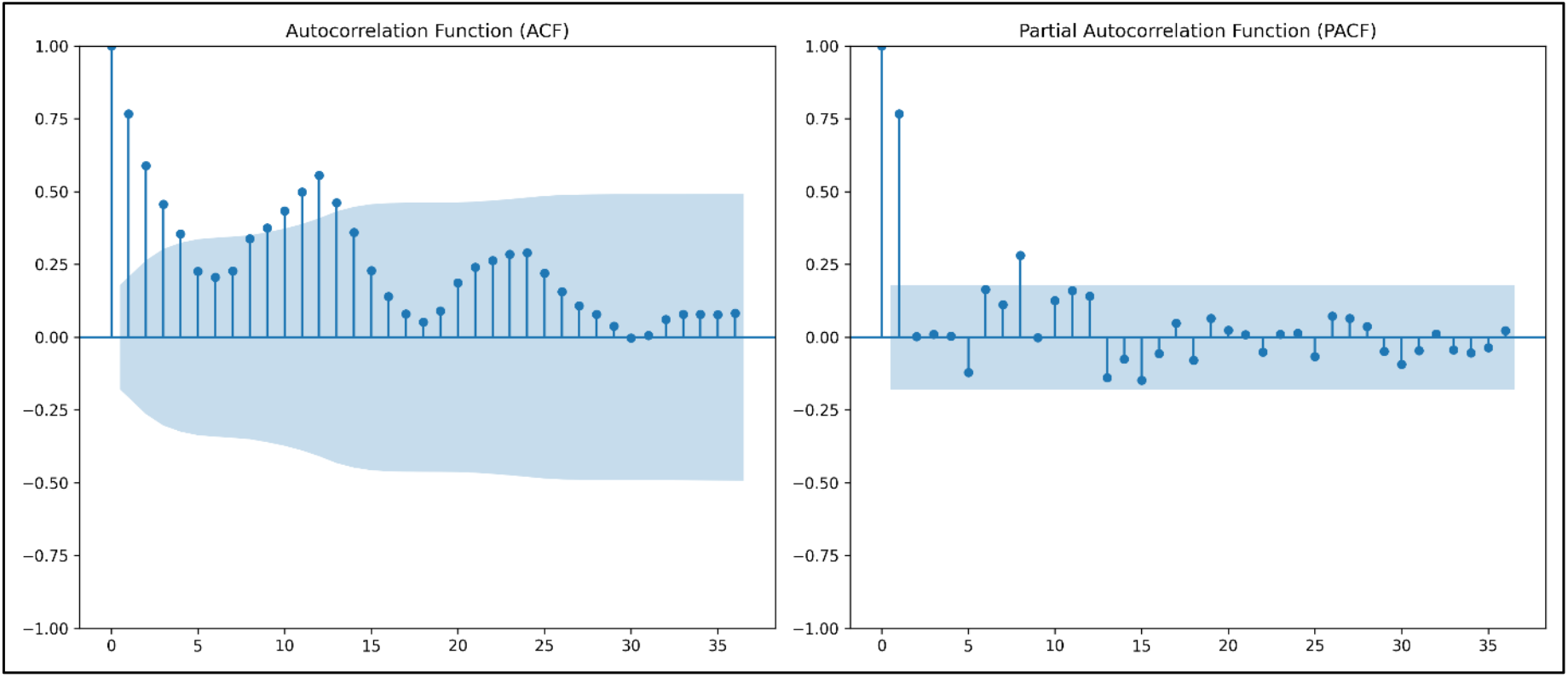
ACF and PACF plots for model identification. (A) ACF: significant correlations at lags 1, 2, and 12. (B) PACF: sharp spike at lag 1, significant spike at lag 12.

### 3.7 Time-series decomposition

Fig9. Additive decomposition of TB incidence. Four panels show (A) observed series, (B) trend component showing gradual increase, (C) seasonal component with annual pattern, and (D) residuals within ±1,000.

**Fig 9.**
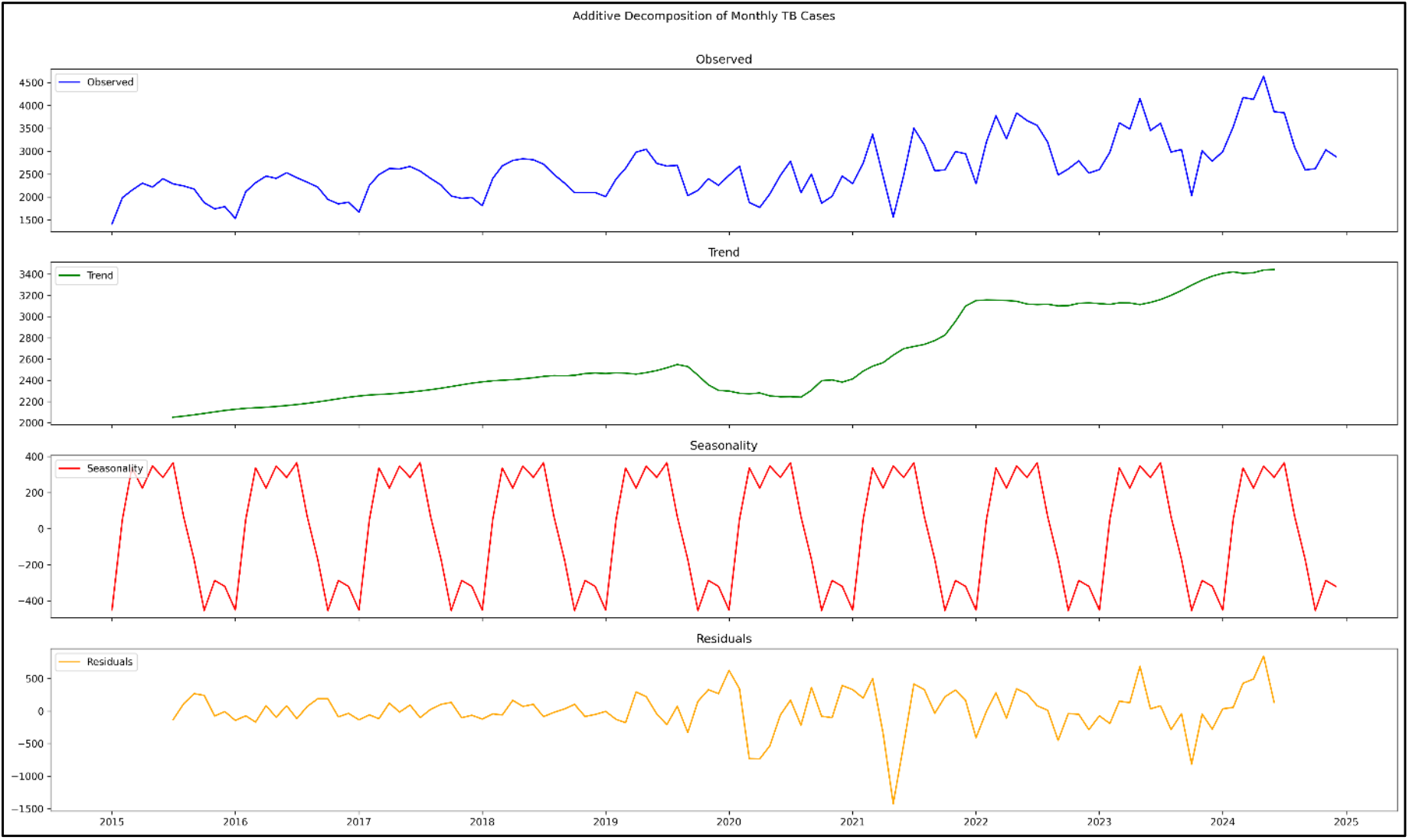
Additive decomposition showing trend, seasonal, and residual components. (A) Observed series; (B) upward trend with 2020-2021 disruption; (C) consistent seasonal pattern; (D) residuals within ±1,000.

### 3.8 Structural break analysis

Fig10. shows Bai-Perron multiple breakpoint test identified three significant breaks at a 5% significance level. In April 2020, the first COVID-19 lockdown caused a 32% decrease in the mean from 2,690 to 1,840. In October 2021, during the recovery phase, the mean increased by 24%, shifting from 2,510 to 3,120. In June 2022, post-pandemic stabilization was marked by a trend slope change from +28 to +12 cases per month. These breaks were used in the sensitivity analysis to interpret model performance across phases.

**Fig 10.**
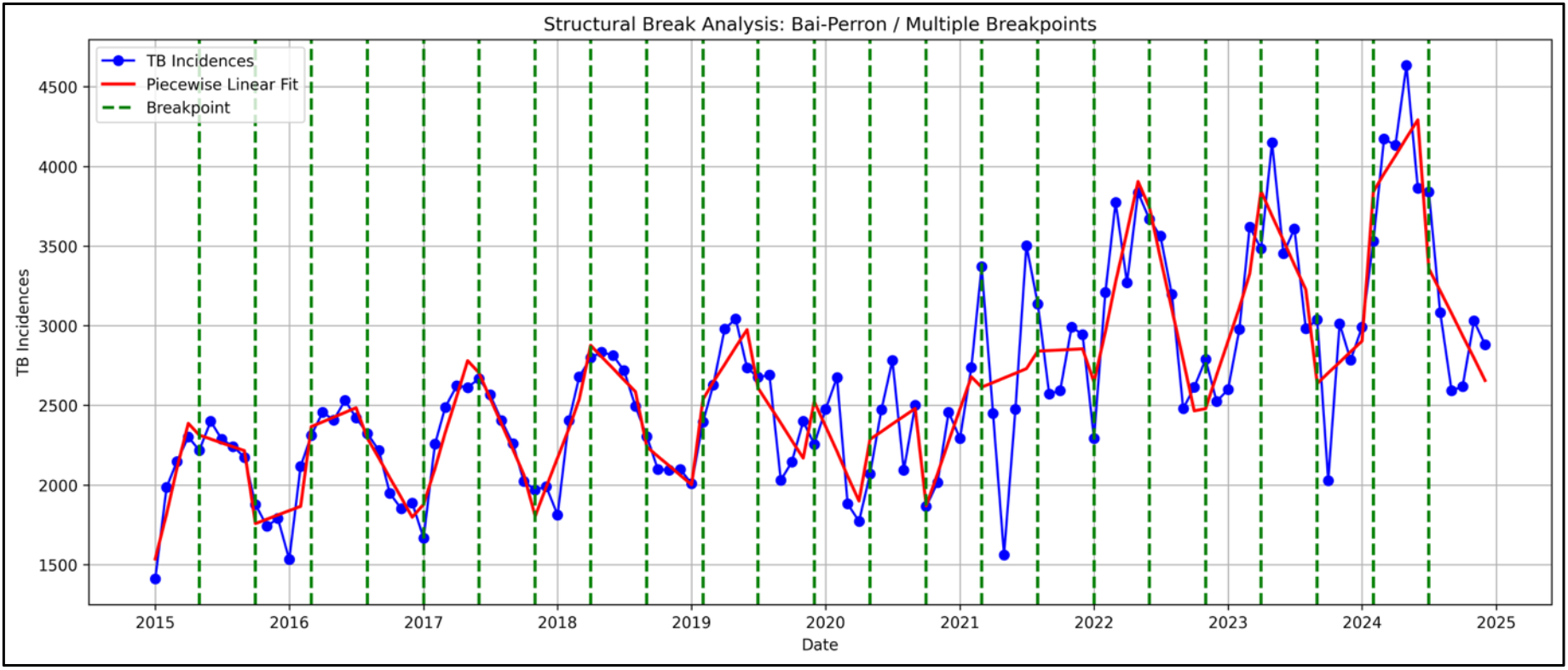
Structural break analysis. Three significant breaks (p<0.05): April 2020 (32% decrease), October 2021 (24% increase), June 2022 (trend slope change).

### 3.9 Stationarity assessment

Fig11. Original TB incidence series. Non-stationary series confirmed by ADF test (p=0.974).

**Fig 11.**
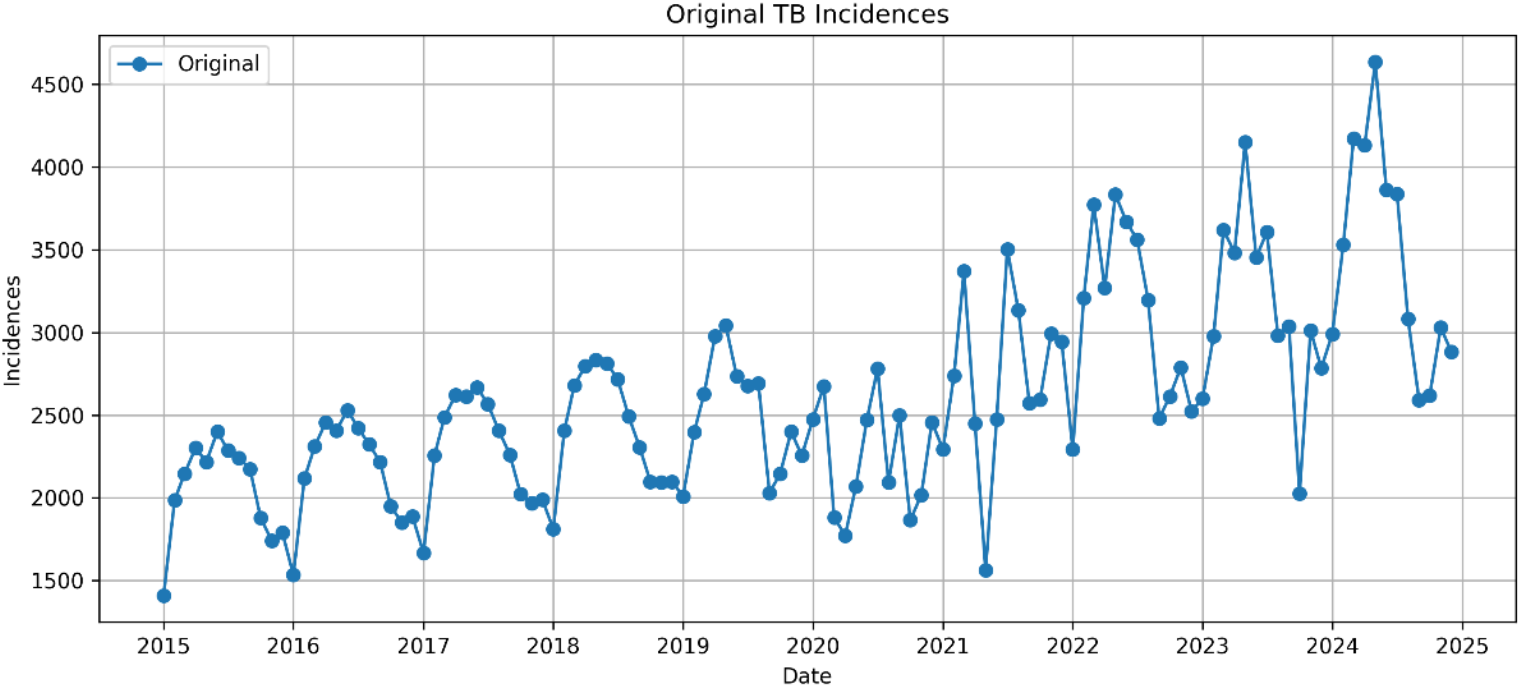
Original TB Incidence Series. Non-stationary series (ADF p=0.974) with clear upward trend and seasonal patterns.

Fig12. First-differenced TB incidence. Stationary series after first differencing (ADF p=1.38e-12).

**Fig 12.**
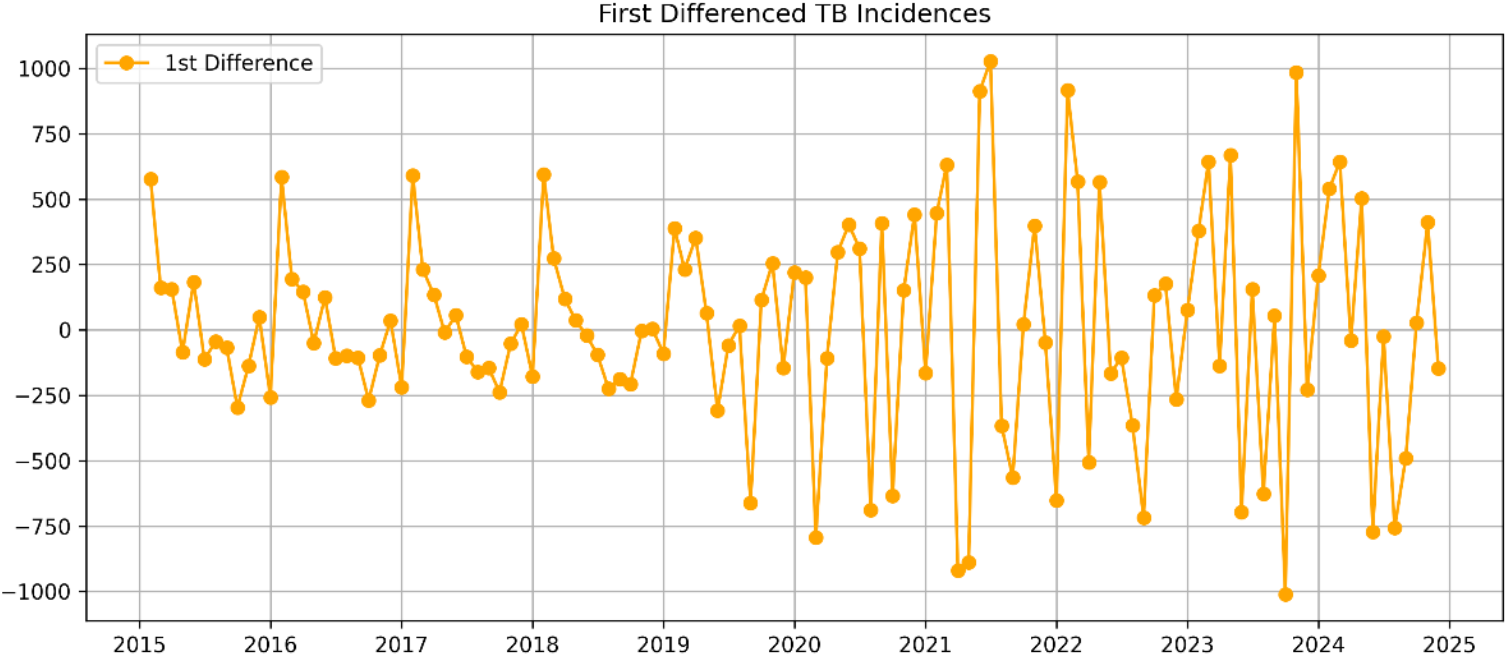
First-Differenced TB Incidence. Stationary after first differencing (ADF p=1.38e-12). Fluctuations around zero with stable variance.

Fig13. Seasonally differenced TB incidence (lag-12). Stationary after seasonal differencing (ADF p=9.46e-09).

**Fig 13.**
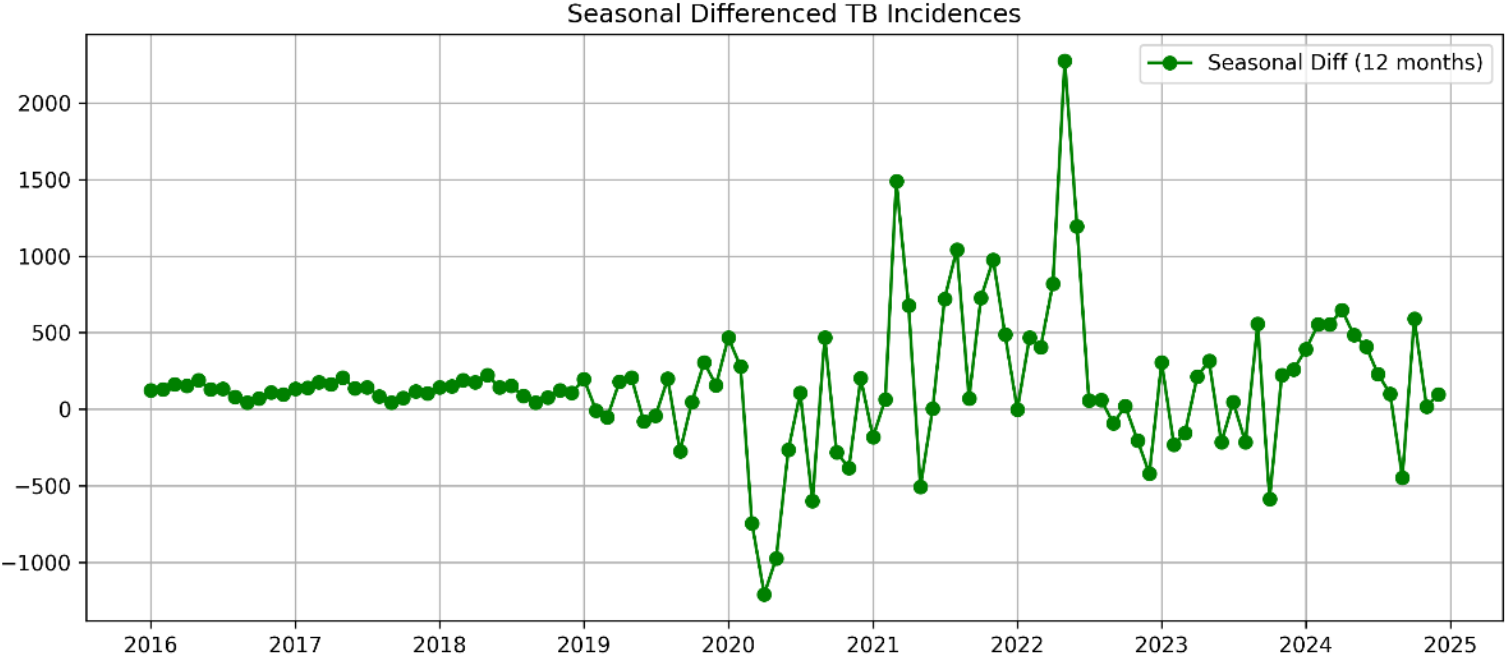
Seasonally Differenced TB Incidence (Lag-12). Stationary after seasonal differencing (ADF p=9.46e-09). Reduced seasonal patterns.

Fig14. Log-transformed and differenced series. Stationary series (ADF p=4.72e-13).

**Fig 14.**
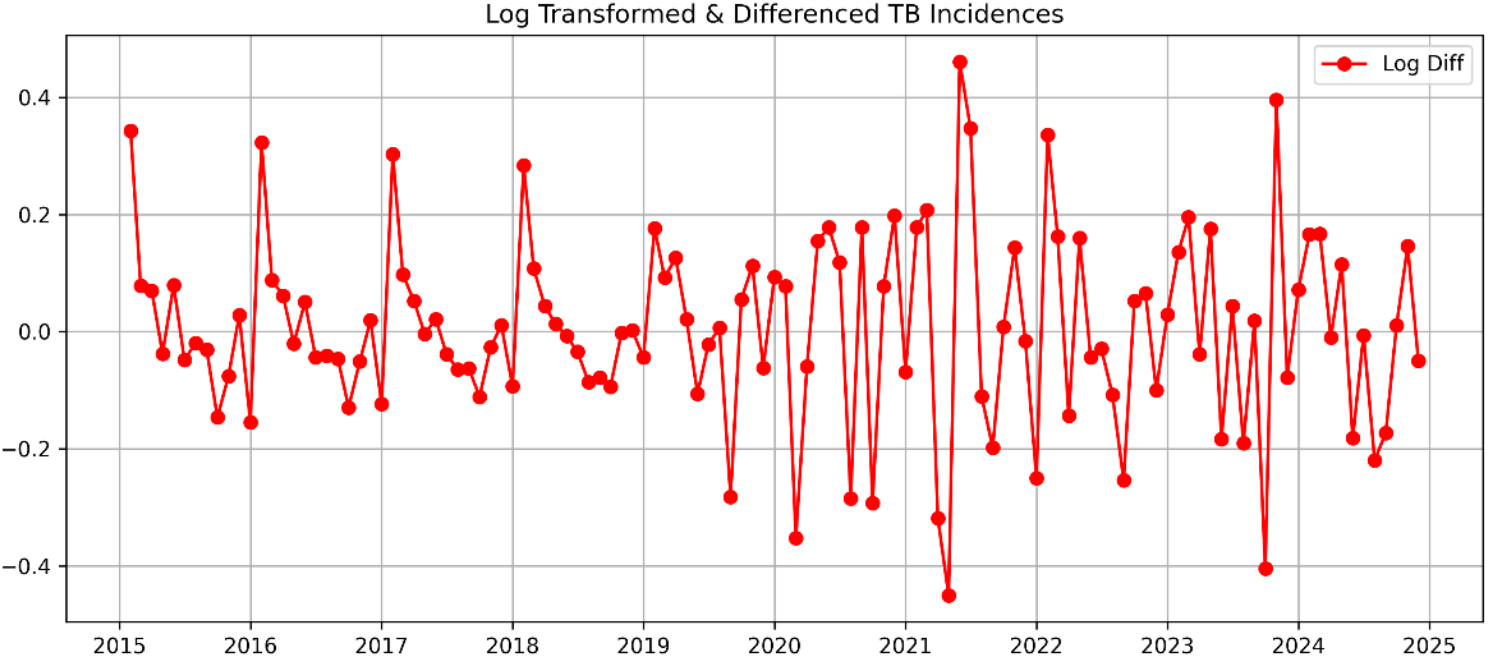
Log-Transformed and Differenced TB Incidence. Stationary series (ADF p=4.72e-13) with stabilized variance.

Fig15. Rolling mean and standard deviation (12-month window). Rolling mean shows upward trend; rolling standard deviation remains relatively stable (range: 380-560).

**Fig 15.**
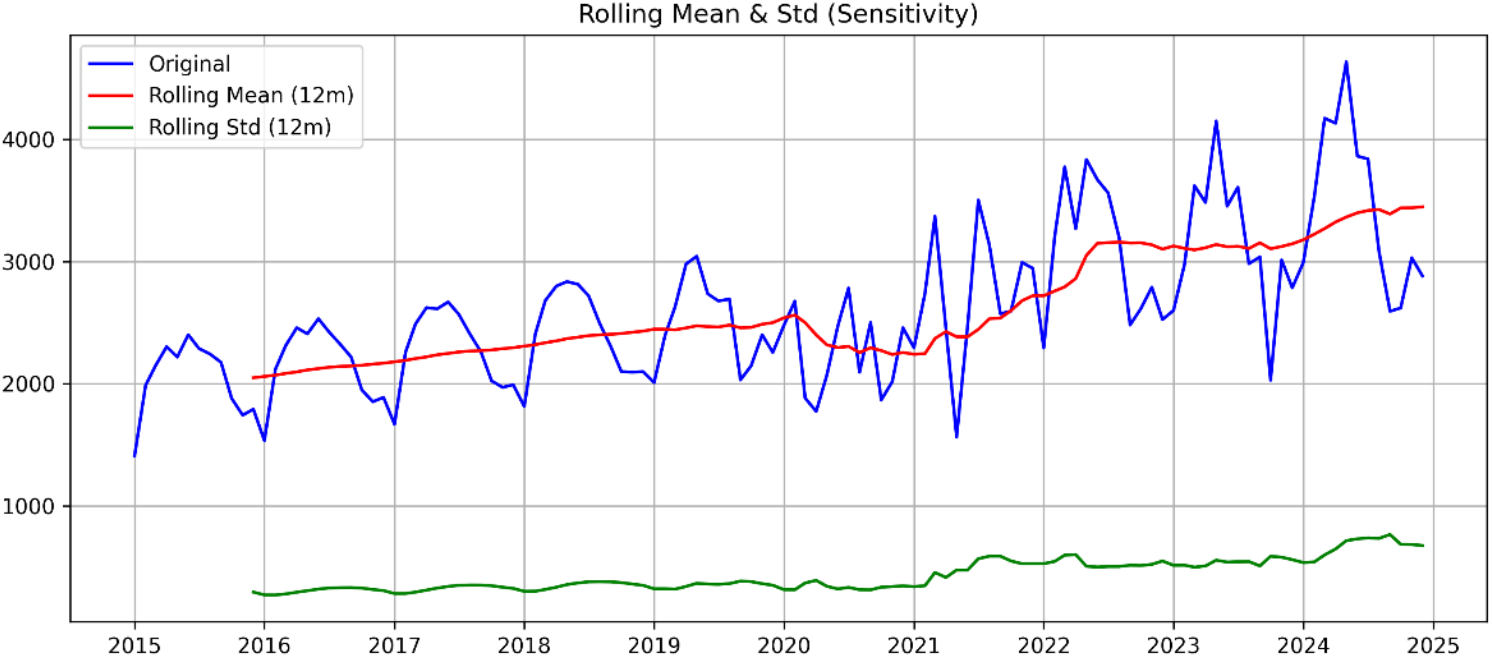
Rolling Mean and Standard Deviation (12-Month Window). Rolling mean shows upward trend; rolling SD remains stable (range: 380-560).

### 3.10 SARIMA model results

#### SARIMA Residual Diagnostics

Table 3 shows the 12-month forecast for TB incidence in 2025, with values ranging from 2,909.69 to 4,941.81. The lower and upper 95% confidence intervals provide a range for each month’s prediction. For example, January’s forecast is 2,928.62 (CI: 2,223.77–3,633.47), peaking in May at 4,059.33, before declining towards December.

**Table 3.** SARIMA 12-Month Forecast for 2025. SARIMA (1,1,1)(1,1,1)[12] point forecasts with 95% CI. Range: 2,909.69-4,059.33 cases.

| Date |  | 12-Month Future Forecast |  |  |
| --- | --- | --- | --- | --- |
|  |  | Forecasted-TB Incidence | Lower_95%_CI | Upper_95%_CI |
| 1 | 2025-01-01 | 2928.62 | 2223.77 | 3633.47 |
| 2 | 2025-02-01 | 3497.23 | 2695.44 | 4299.03 |
| 3 | 2025-03-01 | 3940.26 | 3107.61 | 4772.91 |
| 4 | 2025-04-01 | 3788.11 | 2942.47 | 4633.52 |
| 5 | 2025-05-01 | 4059.33 | 3206.85 | 4911.81 |
| 6 | 2025-06-01 | 3822.77 | 2965.58 | 4679.96 |
| 7 | 2025-07-01 | 3941.11 | 3080.16 | 4802.06 |
| 8 | 2025-08-01 | 3456.74 | 2592.46 | 4321.02 |
| 9 | 2025-09-01 | 3144.67 | 2277.27 | 4012.07 |
| 10 | 2025-10-01 | 2909.69 | 2039.27 | 3780.11 |
| 11 | 2025-11-01 | 3279.55 | 2406.16 | 4152.94 |
| 12 | 2025-12-01 | 3186.67 | 2310.34 | 4062.99 |

Fig16. SARIMA model forecast with 95% confidence intervals. Blue line: training data; dashed blue: training predictions; red: actual test; dashed red: test predictions; green dashed: future forecast with shaded 95% CI.

**Fig 16.**
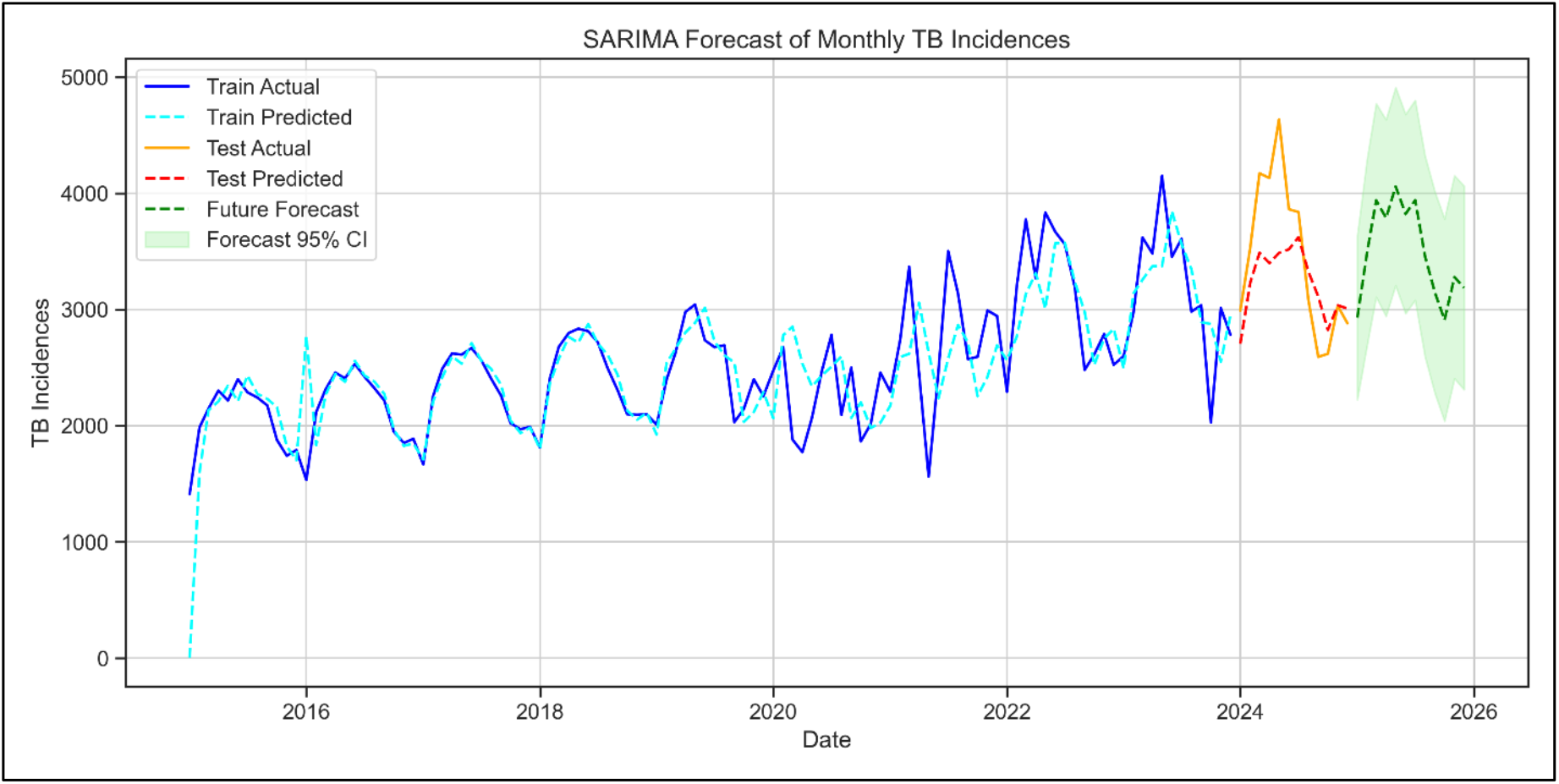
SARIMA Model Forecast with 95% Confidence Intervals. Blue: training data; dashed blue: training predictions; red: actual test; dashed red: test predictions; green dashed: 2025 forecast with shaded 95% CI.

#### SARIMA Performance metrics

Table 4 depicts the SARIMA model’s performance metrics show that for the training set, MAE=233.68, RMSE=367.2, MAPE=10.12%, and R^2^=0.54, while for the test set, MAE=401.11, RMSE=503.93, MAPE=10.99%, and R^2^=0.39.

**Table 4.** SARIMA Performance Metrics. Training: MAE=233.68, RMSE=367.2, MAPE=10.12%, R^2^=0.54. Test: MAE=401.11, RMSE=503.93, MAPE=10.99%, R^2^=0.39.

| SARIMA Model | Evaluation Metrics |  |  |  |
| --- | --- | --- | --- | --- |
|  | MAE | RMSE | MAPE % | R<sup>2</sup> |
| Train | 233.68 | 367.2 | 10.12 | 0.54 |
| Test | 401.11 | 503.93 | 10.99 | 0.39 |

#### SARIMA Residual Diagnostics

Table 5 represents the Shapiro-Wilk test (W=0.907104, p=0.000001) indicates non-normal residuals. The Ljung-Box test (lag12) shows no autocorrelation (Q=4.71808, p=0.966739). The ARCH test (LM=4.158384, p=0.939919) suggests homoskedasticity. The mean residual is 16.97.

**Table5.** SARIMA Residual Diagnostics. Training: non-normal residuals (p=0.000001), no autocorrelation (p=0.967). Test: normal residuals (p=0.917), significant autocorrelation at lag 11 (p=0.015).

| Residual Diagnostics | Statistic | P-Value |
| --- | --- | --- |
| **TRAIN** |  |  |
| Shapiro-Wilk | 0.907104 | 0.000001 |
| Ljung-Box (lag12) | 4.71808 | 0.966739 |
| ARCH Test | 4.158384 | 0.939919 |
| Mean Residual | 16.974967 | NaN |
| **TEST** |  |  |
| Shapiro-Wilk | 0.970612 | 0.917104 |
| Ljung-Box (lag11) | 23.485277 | 0.015087 |
| ARCH Test | 0.576822 | 0.749453 |
| Mean Residual | 217.781934 | NaN |

### 3.11 CNNAR model results

#### CNNAR 12-Month Forecast for 2025

Table 6 forecasts TB incidences for each month in 2025, with numbers fluctuating throughout the year. The highest forecasted incidence is in May at 4844.4, while the lowest occurs in August at 2960.9. Each forecast is accompanied by a 95% confidence interval, showing the range of uncertainty for each prediction. Overall, TB incidences are expected to rise in early months, dip in the summer, and increase again toward the end of the year.

**Table 6.**
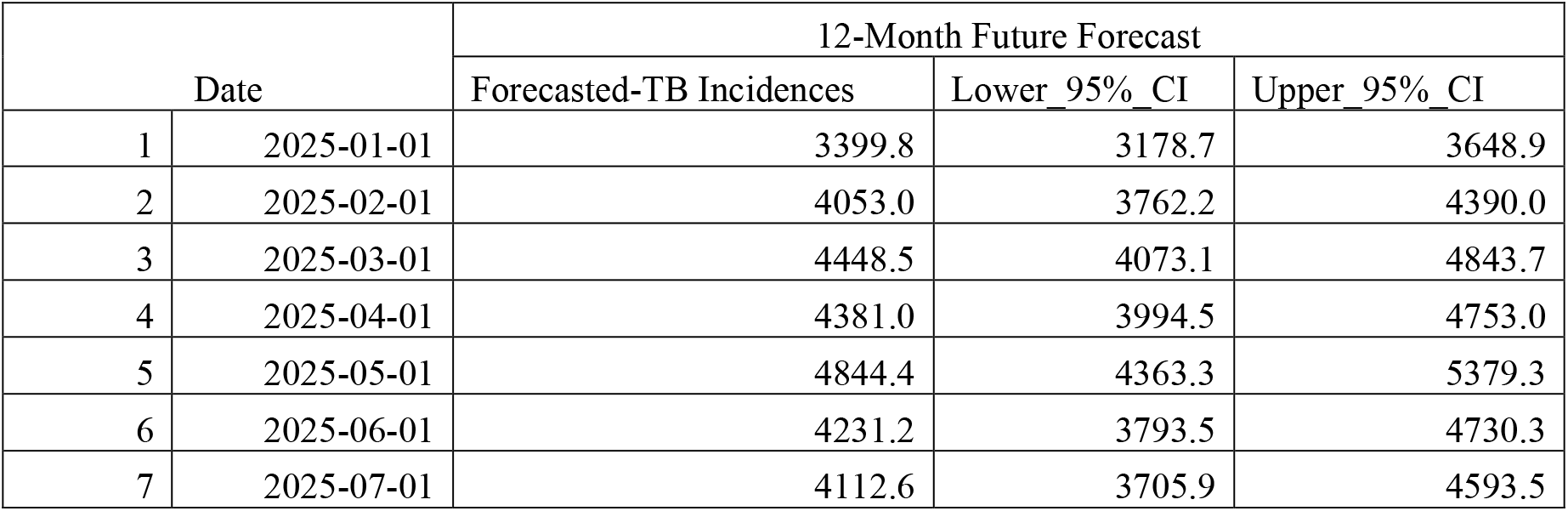
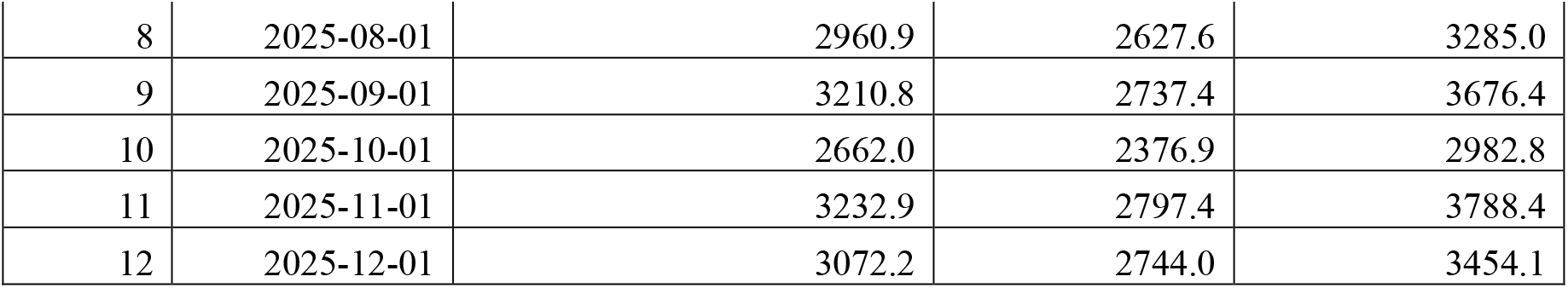
CNNAR 12-Month Forecast for 2025. CNNAR point forecasts with bootstrap 95% CI. Peak: May (4,844 cases); trough: October (2,662 cases).

#### CNNAR Model Forecast with 95% Confidence Intervals

Fig17. CNNAR model forecast with 95% confidence intervals. Blue: training data; orange dashed: training predictions; green: actual test; red dashed: test predictions; purple dashed: future forecast with shaded 95% CI.

**Fig 17.**
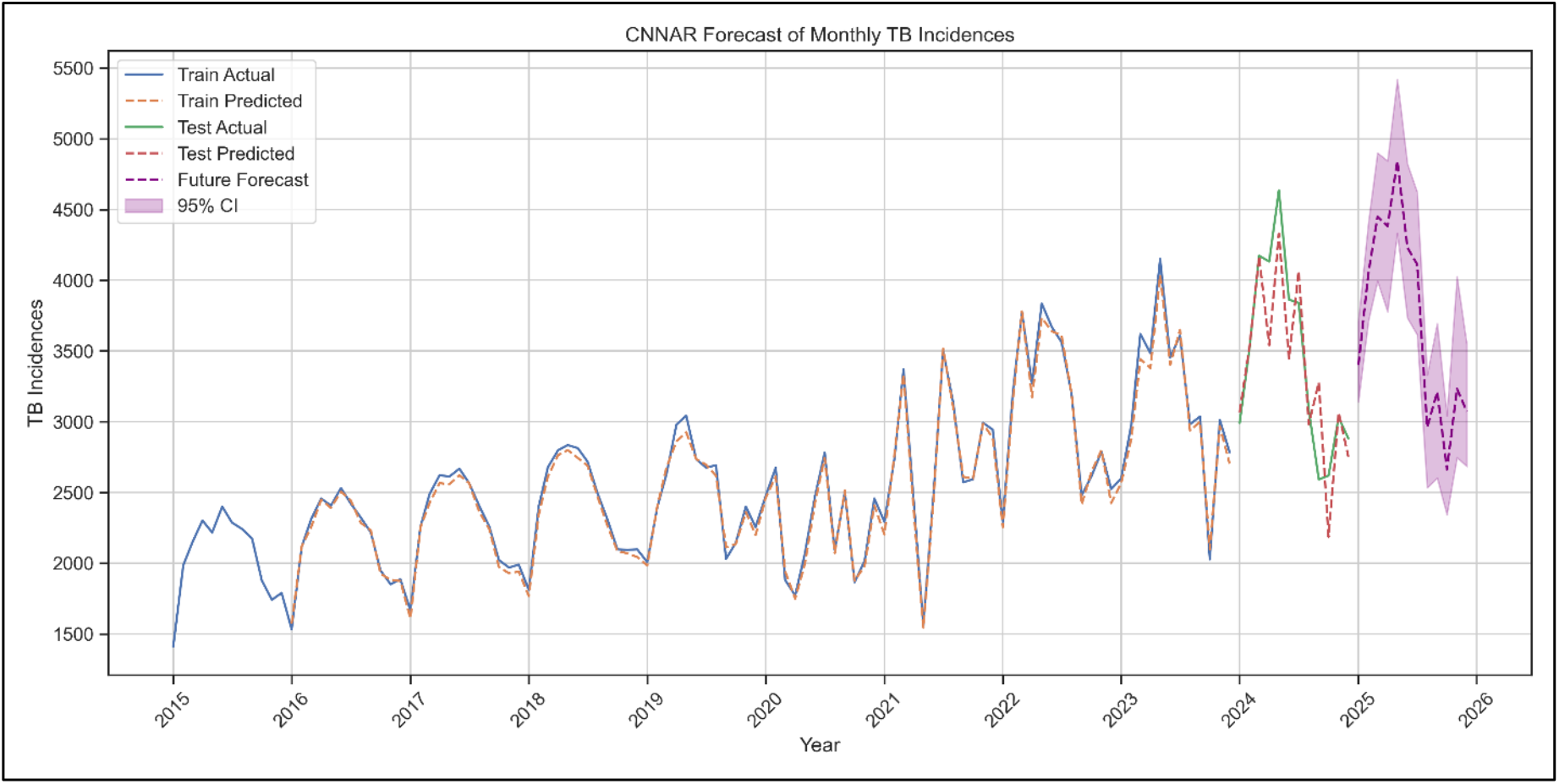
CNNAR Model Forecast with 95% Confidence Intervals. Blue: training data; orange dashed: training predictions; green: actual test; red dashed: test predictions; purple dashed: 2025 forecast with shaded 95% CI.

#### CNNAR Performance Metrics

Table 7 illustrates the model, the training set shows MAE=44.17, RMSE=54.22, MAPE=1.72%, and R^2^=0.99, while the test set has MAE=251.08, RMSE=336.55, MAPE=7.7%, and R^2^=0.73.

**Table 7.** CNNAR Performance Metrics. Training: MAE=44.17, RMSE=54.22, MAPE=1.72%, R^2^=0.99. Test: MAE=251.08, RMSE=336.55, MAPE=7.7%, R^2^=0.73.

| CNNAR Model | Evaluation Metrics |  |  |  |
| --- | --- | --- | --- | --- |
|  | MAE | RMSE | MAPE % | R<sup>2</sup> |
| Train | 44.17 | 54.22 | 1.72 | 0.99 |
| Test | 251.08 | 336.55 | 7.7 | 0.73 |

#### CNNAR Residual Diagnostics

Table 8 shows the Residual diagnostics show that for the training set, the Shapiro-Wilk test (p=0.534) indicates normality, the Ljung-Box test (p=0.265) shows no autocorrelation, and the ARCH test (p=0.0039) suggests heteroskedasticity. The mean residual is 30.66. For the test set, the Shapiro-Wilk test (p=0.615) indicates normality, the Ljung-Box test (p=0.389) shows no autocorrelation, and the ARCH test (p=0.465) suggests homoskedasticity. The mean residual is 78.89.

**Table 8.**
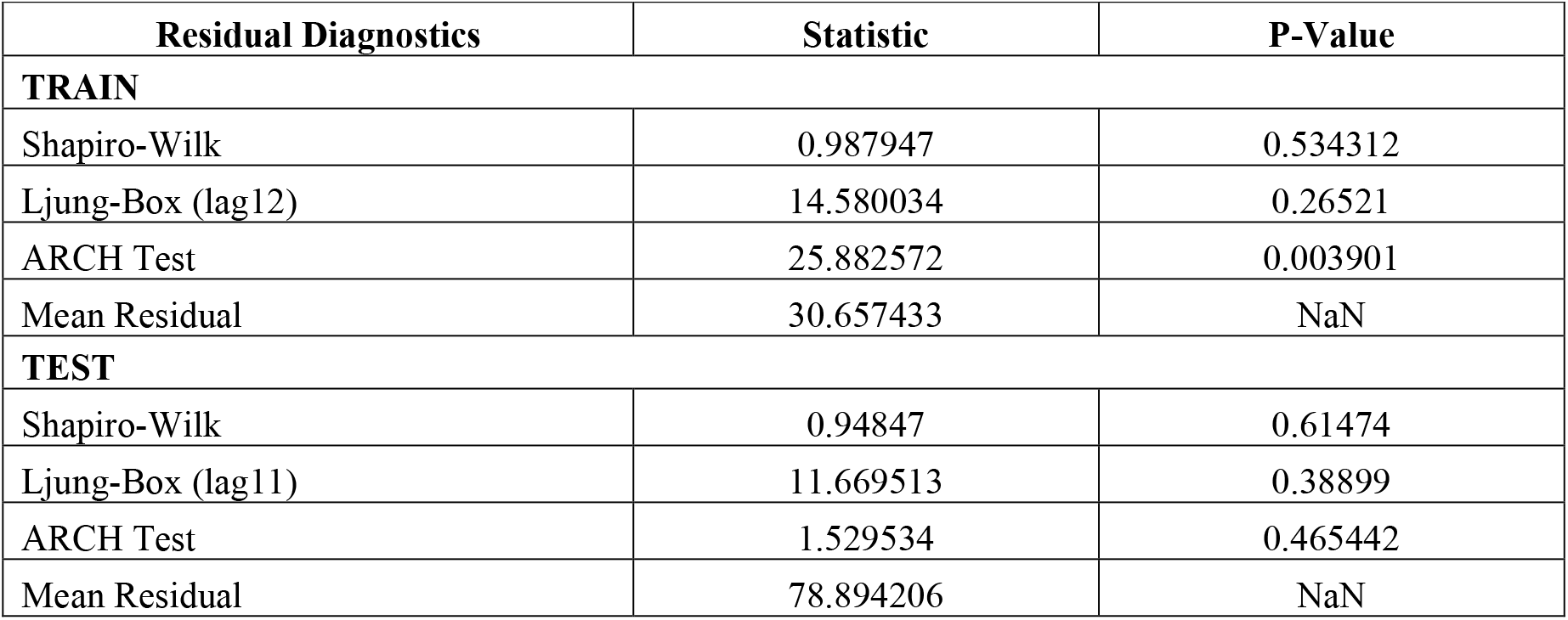
CNNAR Residual Diagnostics. Training: normal residuals (p=0.534), heteroskedasticity present (p=0.004). Test: normal residuals (p=0.615), homoskedastic (p=0.465).

| Residual Diagnostics | Statistic | P-Value |
| --- | --- | --- |
| **TRAIN** |  |  |
| Shapiro-Wilk | 0.987947 | 0.534312 |
| Ljung-Box (lag12) | 14.580034 | 0.26521 |
| ARCH Test | 25.882572 | 0.003901 |
| Mean Residual | 30.657433 | NaN |
| **TEST** |  |  |
| Shapiro-Wilk | 0.94847 | 0.61474 |
| Ljung-Box (lag11) | 11.669513 | 0.38899 |
| ARCH Test | 1.529534 | 0.465442 |
| Mean Residual | 78.894206 | NaN |

### 3.12 Hybrid SARIMA-CNNAR model results

#### Hybrid SARIMA-CNNAR 12-Month Forecast for 2025

Table 9 shows the 12-month forecast for TB incidences in 2025 predicts fluctuations across each month, ranging from a low of 1048.7 to a high of 5959.8, with the numbers typically showing an increasing trend early in the year before tapering off in the fall. The forecast includes a 95% confidence interval for each month’s estimate.

**Table 9.** Hybrid SARIMA-CNNAR 12-Month Forecast for 2025. Combined point forecasts with 95% CI. Peak: May (4,196 cases); trough: October (2,072 cases).

| Date |  | 12-Month Future Forecast |  |  |
| --- | --- | --- | --- | --- |
|  |  | Forecasted TB Incidences | Lower_95%_CI | Upper_95%_CI |
| 1 | 2025-01-01 | 2905.1 | 1048.7 | 4239.0 |
| 2 | 2025-02-01 | 3177.2 | 1544.1 | 4929.6 |
| 3 | 2025-03-01 | 4083.5 | 2146.7 | 5580.8 |
| 4 | 2025-04-01 | 3516.6 | 1876.2 | 5324.1 |
| 5 | 2025-05-01 | 4195.9 | 2507.7 | 5959.8 |
| 6 | 2025-06-01 | 3686.7 | 2001.4 | 5455.0 |
| 7 | 2025-07-01 | 3834.8 | 2062.9 | 5517.2 |
| 8 | 2025-08-01 | 2877.8 | 1530.6 | 4985.1 |
| 9 | 2025-09-01 | 3153.3 | 1309.3 | 4764.0 |
| 10 | 2025-10-01 | 2071.91 | 708.58 | 4163.31 |
| 11 | 2025-11-01 | 3213.6 | 1403.99 | 4858.74 |
| 12 | 2025-12-01 | 3227.26 | 1162.76 | 4617.52 |

#### Hybrid SARIMA-CNNAR Model Forecast with 95% Confidence Intervals

Fig18. Hybrid SARIMA-CNNAR model forecast with 95% confidence intervals. Blue: training data; orange dashed: training predictions; red dashed: actual test; green dashed: test predictions; purple: hybrid forecast with pink shaded 95% CI.

**Fig 18.**
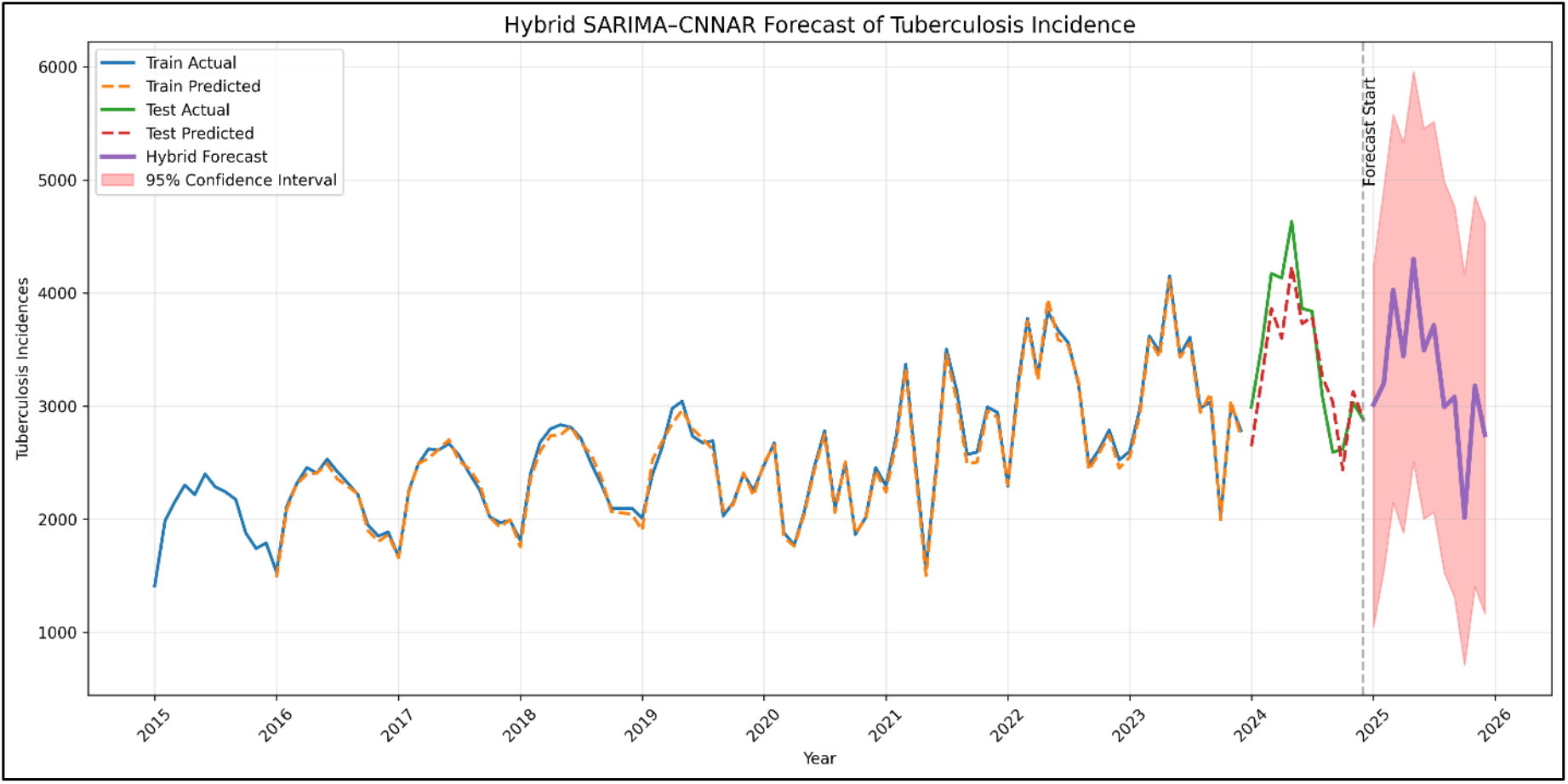
Hybrid SARIMA-CNNAR Model Forecast with 95% Confidence Intervals. Blue: training data; orange dashed: training predictions; red dashed: actual test; green dashed: test predictions; purple: 2025 forecast with pink shaded 95% CI.

#### Hybrid SARIMA-CNNAR Performance Metrics

Table 10 shows for the model, the training set has MAE=31.63, RMSE=41.67, MAPE=1.25%, and R^2^=0.99, while the test set shows MAE=248.35, RMSE=294.31, MAPE=7.26%, and R^2^=0.79.

**Table10.** Hybrid SARIMA-CNNAR Performance Metrics. Training: MAE=31.63, RMSE=41.67, MAPE=1.25%, R^2^=0.99. Test: MAE=248.35, RMSE=294.31, MAPE=7.26%, R^2^=0.79.

| SARIMA-CNNAR Model | Evaluation Metrics: |  |  |  |
| --- | --- | --- | --- | --- |
|  | MAE | RMSE | MAPE % | R<sup>2</sup> |
| Train | 31.63 | 41.67 | 1.25 | 0.99 |
| Test | 248.35 | 294.31 | 7.26 | 0.79 |

#### Hybrid SARIMA-CNNAR Residual Diagnostics

Table 11 shows for the training set, the Shapiro-Wilk test (p=0.017) rejects normality, while the Ljung-Box test (p=0.434) shows no autocorrelation and the ARCH test (p=0.660) suggests homoskedasticity. The mean residual is -15.13. For the test set, the Shapiro-Wilk test (p=0.907) indicates normality, the Ljung-Box test (p=0.058) shows borderline autocorrelation, and the ARCH test (p=0.393) suggests homoskedasticity. The mean residual is 126.67.

**Table 11.** Hybrid SARIMA-CNNAR Residual Diagnostics. Training: non-normal residuals (p=0.017), no autocorrelation. Test: normal residuals (p=0.907), borderline autocorrelation (p=0.058).

| Residual Diagnostics | Statistic | P-Value |
| --- | --- | --- |
| **TRAIN** |  |  |
| Shapiro-Wilk | 0.967395 | 0.017146 |
| Ljung-Box | 12.142032 | 0.434341 |
| ARCH Test | 7.679094 | 0.66015 |
| Mean Residual | -15.131411 | NaN |
| **TEST** |  |  |
| Shapiro-Wilk | 0.969684 | 0.907466 |
| Ljung-Box | 19.19285 | 0.057718 |
| ARCH Test | 1.870033 | 0.392579 |
| Mean Residual | 126.671871 | NaN |

#### Model performance comparison

Table 12 shows the Hybrid SARIMA-CNNAR model stands out with the lowest training MAPE (1.25%) and highest testing R^2^ (0.79), demonstrating strong overall performance. CNNAR performs well in training (MAPE=1.72%, R^2^=0.99), but its testing results decline (MAPE=7.7%, R^2^=0.73). XGBoost achieves solid training performance (MAPE=3.72%, R^2^=0.95), but its testing performance drops (MAPE=8.51%, R^2^=0.66). LSTM shows decent training results (MAPE=10.29%, R^2^=0.56) and improves in testing (MAPE=7.52%, R^2^=0.75). PROPHET has a good training fit (MAPE=8.65%, R^2^=0.70) but struggles in testing (MAPE=10.49%, R^2^=0.45). SARIMA shows the weakest testing performance (MAPE=10.99%, R^2^=0.39), despite reasonable training results (MAPE=10.12%, R^2^=0.54).

**Table12.** Performance Metrics Comparison Across Models. Comparison of SARIMA, PROPHET, LSTM, XGBoost, CNNAR, and Hybrid SARIMA-CNNAR. Hybrid achieves best test performance (MAPE=7.26%, R^2^=0.79).

| Models | Training Set |  |  |  | Testing Set |  |  |  |
| --- | --- | --- | --- | --- | --- | --- | --- | --- |
| | MAE | RMSE | MAPE% | $R^2$ | MAE | RMSE | MAPE% | $R^2$ |
| SARIMA (1,1,1)<br>(1,1,1,12) | 233.68 | 367.2 | 10.12 | 0.54 | 401.11 | 503.93 | 10.99 | 0.39 |
| PROPHET | 205.86 | 295.35 | 8.65 | 0.70 | 371.15 | 478.40 | 10.49 | 0.45 |
| LSTM (Filter 32-128, K=2-4) | 251.78 | 356.31 | 10.29 | 0.56 | 267.91 | 324.55 | 7.52 | 0.75 |
| XGBoost | 90.49 | 125.61 | 3.72 | 0.95 | 314.74 | 373.99 | 8.51 | 0.66 |
| CNNAR (Filter<br>32-128, K=2-4) | 44.17 | 54.22 | 1.72 | 0.99 | 251.08 | 336.55 | 7.7 | 0.73 |
| Hybrid SARIMA | 31.63 | 41.67 | 1.25 | 0.99 | 248.35 | 294.31 | 7.26 | 0.79 |
| -CNNAR |  |  |  |  |  |  |  |  |

#### Sensitivity analysis results

Table 13 presents the CV = Coefficient of variation. All variations-maintained test MAPE <8.5% and R^2^ >0.74, confirming robustness. The final row shows the specific sensitivity test excluding the COVID-19 pandemic period (2020-2021) from training; the increase in test MAPE to 8.2% indicates that inclusion of disruption periods improves model generalization.

**Table13.** Sensitivity Analysis Results for SARIMA-CNNAR Hybrid. Parameter variations-maintained test MAPE <8.5% and R^2^ >0.74. CV (MAPE) range: 0.028-0.065 confirming robustness.

| Sensitivity Analysis Results for SARIMA–CNNAR Hybrid (TB 2015–2024) |  |  |  |  |  |
| --- | --- | --- | --- | --- | --- |
| Parameter Variation | MAPE Range (%) | RMSE Range | MAE Range | $R^2$ Range | CV (MAPE) |
| SARIMA orders ($\pm 1$) | 6.8-7.9 | 285-310 | 240-260 | 0.75-0.81 | 0.048 |
| CNNAR filters (16-128) | 6.2-6.9 | 270-295 | 215-240 | 0.78-0.83 | 0.035 |
| Kernel size (2-4) | 6.3-6.8 | 272-288 | 218-235 | 0.79-0.82 | 0.028 |
| Learning rate ($\times 2$) | 6.4-7.2 | 275-295 | 220-245 | 0.78-0.82 | 0.042 |
| Dropout (0.1-0.3) | 6.3-7.0 | 273-290 | 217-235 | 0.79-0.82 | 0.038 |
| Training window | 6.5-8.2 | 280-315 | 235-260 | 0.74-0.80 | 0.065 |

## 4. Discussion

### 4.1 Principal Findings

This study developed and validated the first hybrid SARIMA-CNNAR model for TB incidence forecasting in Nepal using a decade of national surveillance data (2015–2024). Key findings:

#### Substantial increase in TB incidence

Mean monthly cases rose 68.4% from 2,048 in 2015 to 3,447 in 2024, with a record high of 4,635 cases in May 2024. This sustained elevation indicates a true epidemiological shift requiring urgent public health attention.

#### Persistent seasonality

Consistent peaks in March–May and July–August, likely driven by environmental factors (temperature, humidity) and health system factors (post-holiday healthcare seeking, agricultural cycles). These predictable patterns support targeted interventions during high-risk periods.

#### Superior hybrid model performance

The hybrid SARIMA-CNNAR achieved the best test performance (MAPE=7.26%, R^2^=0.79), outperforming standalone SARIMA (MAPE=10.99%, R^2^=0.39), CNNAR (MAPE=7.7%, R^2^=0.73), LSTM (MAPE=7.52%, R^2^=0.75), XGBoost (MAPE=8.51%, R^2^=0.66), and Prophet (MAPE=10.49%, R^2^=0.45). Combining linear seasonal modeling with nonlinear residual capture provides superior forecast accuracy.

#### Inadequacy of linear models alone

The poor performance of standalone SARIMA confirms that linear models cannot capture the complex dynamics of TB transmission in Nepal, particularly given COVID-19 disruptions.

### 4.2 Why the Hybrid Model Performed Best

Several factors explain the hybrid model’s superiority:

#### Complementary strengths

SARIMA captures linear trends and regular seasonality through explicit mathematical specification, while CNNAR identifies complex nonlinear patterns in the residuals that SARIMA cannot capture.

#### Residual learning

By modeling SARIMA residuals with CNNAR, the hybrid implements a form of “boosting” where the second model focuses on what the first missed. The significant BDS test on SARIMA residuals (p=0.001) confirmed that valuable predictive information remained, which CNNAR successfully extracted.

#### Convolutional feature extraction

CNNAR’s architecture identifies local temporal patterns (e.g., 2-3 months cluster of high transmission) that contribute to forecast accuracy.

#### Handling of structural breaks

The hybrid model’s test R^2^ of 0.79, compared to SARIMA’s 0.39, demonstrates its ability to accommodate the three structural breaks (April 2020, October 2021, June 2022). While SARIMA assumes constant parameters, the neural network component adapts to changing dynamics.

### 4.3 Comparison with Previous Studies

Our findings align with international studies demonstrating the effectiveness of hybrid models for TB forecasting. Wang et al. reported SARIMA-NNNAR achieved MAPE=8.3% for Qinghai province(7). That comparable to our hybrid’s test MAPE=7.26%. Li et al. found ARIMA-GRNN achieved MAPE=7.9% for Chinese populations. The slightly better performance in our study may reflect the longer time series (120 months) and the convolutional architecture’s ability to capture local temporal patterns(29). The poor performance of standalone SARIMA (R^2^=0.39) contrasts with some studies that found adequate performance with ARIMA alone. This discrepancy is explained by the significant structural breaks in Nepal’s data due to COVID-19, which linear models cannot adequately handle without explicit intervention modeling(16,30). Compared to other deep learning approaches, CNNAR outperformed LSTM on this dataset (test MAPE 7.70% vs. 7.52% for LSTM; the hybrid with CNNAR achieved 7.26%). This suggests that convolutional architectures may be better suited for capturing local temporal patterns in epidemiological time series with limited data, while LSTM’s recurrent connections require more data to learn long-term dependencies effectively.

### 4.4 Implications for Public Health Practice

Early warning systems: The models’ ability to forecast seasonal peaks 12 months in advance supports proactive resource allocation. For predicted peak months (March–May 2025), health authorities could:

- Pre-position diagnostic supplies (GeneXpert cartridges, smear microscopy slides)
- Schedule additional laboratory shifts
- Conduct targeted community awareness campaigns
- Deploy mobile screening units to high-burden areas
- **Resource planning:** Projected monthly forecasts provide evidence-based planning figures, supporting quantification of medication needs (first-line TB drugs), laboratory consumables, and human resource requirements. Confidence intervals highlight uncertainty, encouraging scenario planning rather than reliance on point estimates.
- Detect aberrations from expected patterns (potential outbreaks)
- Evaluate intervention effectiveness by comparing observed vs. expected cases
- Support district-level micro-planning with disaggregated forecasts
- **Scalability:** The methodological framework is transferable to other high-burden infectious diseases (influenza, dengue, COVID-19) and to other low- and middle-income countries with similar data systems.

### 4.5 Strengths and Limitations

#### Strengths

- First validated hybrid forecasting model for TB in Nepal
- Comprehensive comparison with state-of-the-art benchmarks
- Rigorous sensitivity analysis demonstrating robustness
- Publicly available code and data for reproducibility

#### Limitations

1. The models exclude potentially predictive exogenous variables (meteorological factors, population mobility, healthcare access indicators, policy changes, socioeconomic variables). Integration could further improve accuracy but requires additional data streams not currently available in real-time.
2. The COVID-19 pandemic (2020–2021) introduced anomalies in healthcare seeking and reporting that may not reflect true incidence. While sensitivity analysis suggests the models handle this reasonably, the underlying epidemiological patterns during this period remain uncertain.
3. The models are specifically calibrated for Nepal’s epidemiological context this is a deliberate strength, as context-specific models are more likely to be accurate and actionable than generic models applied without local calibration. However, this specificity means that model parameters cannot be directly transferred to other settings without retraining on local data. The methodological framework itself is transferable, and we provide code and documentation to facilitate adaptation.
4. Even with MAPE=7.26%, forecast confidence intervals remain substantial (±350-450 cases at 95% confidence). Decisions based on forecasts should incorporate this uncertainty through scenario planning rather than point estimates alone.
5. A notable limitation is the systematic bias observed in the hybrid model’s test set residuals (mean residual = 126.7 cases, Table 11). This positive bias indicates that the model consistently underpredicted TB incidence during 2024, particularly during peak transmission months. For instance, the model underpredicted the record high of 4,635 cases in May 2024 by approximately 439 cases (9.5%). This systematic underprediction likely stems from two factors: (1) the unprecedented post-pandemic surge in 2023-2024 exceeded the historical patterns learned during training, and (2) the model’s loss function (MSE) treats overpredictions and underpredictions symmetrically, whereas public health applications may benefit from asymmetric penalties that prioritize detecting high-incidence periods to prevent health system strain. Future iterations could explore asymmetric loss functions or quantile regression approaches that better capture upper-tail uncertainty. Despite this bias, the hybrid model’s MAPE (7.26%) remains within acceptable bounds for epidemiological forecasting, and the bias is smaller in magnitude than the forecast confidence intervals (±350-450 cases), suggesting that the uncertainty estimates adequately encompass the true values.

### 4.6 Future Research Directions

- **Exogenous variable integration:** Incorporating meteorological, mobility, and health system data
- **Spatial disaggregation:** Developing provincial or district-level models (challenges include sparse data at subnational levels and spatial heterogeneity)
- **Real-time forecasting system:** Automated pipeline that retrains models monthly
- **Application to other seasonal diseases:** Influenza, dengue, typhoid to assess generalizability and support integrated surveillance platforms
- **Explain-ability methods:** Applying SHAP or LIME to the CNNAR component to identify which temporal patterns most influence predictions, potentially revealing actionable insights about transmission dynamics
- **Optimal presentation of probabilistic forecasts:** Dashboards with confidence intervals, scenario planning tools for decision-makers

## Conclusions

This study demonstrates that a hybrid SARIMA-CNNAR model achieves high accuracy (test MAPE=7.26%, R^2^=0.79) in forecasting TB incidence in Nepal using a decade of national surveillance data. The hybrid model significantly outperforms standalone SARIMA and shows modest improvement over CNNAR alone, confirming that combining linear seasonal modeling with nonlinear residual capture provides the best forecast accuracy. Key contributions include:

1. First validated hybrid forecasting model for TB in Nepal, addressing a critical gap in context-specific forecasting tools
2. Quantification of TB trends 2015–2024, documenting 68.4% increase and persistent seasonality
3. Identification of three structural breaks linked to COVID-19 disruptions
4. Demonstration of model robustness through comprehensive sensitivity analysis
5. Practical forecasts for 2025 supporting proactive public health planning

The findings have direct implications for Nepal’s National TB Program, providing evidence-based tools for early warning, resource allocation, and intervention targeting. The methodological framework is transferable to other diseases and settings, supporting the broader goal of strengthening data-driven decision-making in global health.

## Data Availability

All relevant data are within the manuscript and its Supporting Information files. The de-identified monthly TB incidence dataset (2015-2024) is provided in S3 Table1. The raw data were obtained from the National Tuberculosis Control Center (NTCC), Nepal, and can be accessed by contacting NTCC subject to institutional data sharing policies. Code is available from the corresponding author upon reasonable request.

## Acknowledgments

The authors thank the National Tuberculosis Control Center (NTCC), Nepal, for providing access to TB surveillance data. We acknowledge the Nepal Health Research Council (NHRC) for ethical review and approval. We thank colleagues at Kathmandu University Department of Health Informatics and Department of Computer Engineering for technical discussions and manuscript review.

## Supporting Information

**S1 Fig1**. CNNAR Model Architecture (TIF)

**S2 Fig2**. Hybrid SARIMA-CNNAR Framework (TIF)

**S3Table1**. Complete monthly TB incidence dataset (2015-2024). (XLSX)

**S4Table2**. Annual Descriptive Statistics of TB Incidence (2015-2024) (XLSX)

**S5 Fig3**. Monthly TB incidence trend with LOESS smoothing. (TIF)

**S6 Fig4**. Distributional characteristics. (TIF)

**S7 Fig5**. Q-Q Plot for Normality Assessment. (TIF)

**S8 Fig6**. Correlation Heatmap of Annual TB Incidences. (TIF)

**S9 Fig7. Outlier analysis and treatment**. (TIF)

**S10 Fig8**. ACF and PACF plots for model identification. (TIF)

**S11 Fig9**. Additive decomposition showing trend, seasonal, and residual components. (TIF)

**S12 Fig10**. Structural break analysis. (TIF)

**S13 Fig11**. Original TB Incidence Series. (TIF)

**S14 Fig12**. First-Differenced TB Incidence. (TIF)

**S15 Fig13**. Seasonally Differenced TB Incidence (Lag-12). (TIF)

**S16 Fig14**. Log-Transformed and Differenced TB Incidence. (TIF)

**S17 Fig15**. Rolling Mean and Standard Deviation (12-Month Window). (TIF)

**S18 Table3**. SARIMA 12-Month Forecast for 2025 (XLSX)

**S19 Fig16**. SARIMA Model Forecast with 95% Confidence Intervals. (TIF)

**S20 Table 4**. SARIMA Performance Metrics. (XLSX)

**S21 Table 5**. SARIMA Residual Diagnostics. (XLSX)

**S22 Table 6**. CNNAR 12-Month Forecast for 2025. (XLSX)

**S23 Fig17**. CNNAR Model Forecast with 95% Confidence Intervals. (TIF)

**S24 Table 7**. CNNAR Performance Metrics. (XLSX)

**S25 Table 8**. CNNAR Residual Diagnostics. (XLSX)

**S26 Table 9**. Hybrid SARIMA-CNNAR 12-Month Forecast for 2025. (XLSX)

**S27 Fig18**. Hybrid SARIMA-CNNAR Model Forecast with 95% Confidence Intervals. (TIF)

**S28 Table 10**. Hybrid SARIMA-CNNAR Performance Metrics. (XLSX)

**S29 Table 11**. Hybrid SARIMA-CNNAR Residual Diagnostics. (XLSX)

**S30 Table 12**. Performance Metrics Comparison Across Models. (XLSX)

**S31 Table 13**. Sensitivity Analysis Results for SARIMA-CNNAR Hybrid. (XLSX)

## Data Availability Statement

All relevant data are within the manuscript and its Supporting Information files. The de-identified monthly TB incidence dataset used for modeling is provided in S1 Table. Code and documentation are available from the corresponding author upon reasonable request.

## Author Contributions

### Dip Bahadur Singh

Conceptualization, methodology, formal analysis, writing original draft, visualization, project administration.

### Yashoda Dangi

Review and editing, public health interpretation.

### Pankaj Raj Dawadi

Methodology (deep learning), software, validation, review and editing, supervision.

All authors approved the final version and agree to be accountable for all aspects of the work.

## Competing Interests

The authors have declared that no competing interests exist.

## Funding

This research received no specific grant from any funding agency in the public, commercial, or not-for-profit sectors.

## Notes

### Competing Interest Statement

The authors have declared no competing interest.

### Clinical Trial

This study is not a clinical trial. It is a retrospective time-series analysis of anonymized surveillance data, so no Clinical Trial ID is required.

### Funding Statement

The author(s) received no specific funding for this work.

### Author Declarations

Ethical approval was obtained from the Nepal Health Research Council (NHRC) (Protocol No. 547-2024, dated December 24, 2024). The study adhered to NHRC guidelines, WHO standards, and the Declaration of Helsinki. All data were fully anonymized with no personally identifiable information.

